# Deep learning on electronic medical records identifies distinct subphenotypes of diabetic kidney disease driven by genetic variations in the *Rho* pathway

**DOI:** 10.1101/2023.09.06.23295120

**Authors:** Ishan Paranjpe, Xuan Wang, Nanditha Anandakrishnan, Jonathan C. Haydak, Tielman Van Vleck, Stefanie DeFronzo, Zhengzhe Li, Anthony Mendoza, Ruijie Liu, Jia Fu, Iain Forrest, Weibin Zhou, Kyung Lee, Ross O’Hagan, Sergio Dellepiane, Kartikeya M. Menon, Faris Gulamali, Samir Kamat, Gabriele Luca Gusella, Alexander W. Charney, Ira Hofer, Judy H. Cho, Ron Do, Benjamin S Glicksberg, John C. He, Girish N. Nadkarni, Evren U. Azeloglu

**Author notes:** Contributed equally, joint first authors (Ishan Paranjpe, Xuan Wang, Nanditha Anandakrishnan). Supervised jointly; Corresponding Authors: Evren U. Azeloglu, PhD Associate Professor of Medicine, Barbara T Murphy Division of Nephrology Icahn School of Medicine at Mount Sinai Voice; Girish N. Nadkarni, MD, MPH, Irene and Dr. Arthur Fishberg Professor of Medicine, Barbara T Murphy Division of Nephrology, Chief, Division of Data Driven, Icahn School of Medicine at Mount Sinai Voice; John Cijiang He, MD, PhD, Irene and Dr. Arthur Fishberg Professor of Medicine, Chief, Barbara T Murphy Division of Nephrology Icahn School of Medicine at Mount Sinai, Voice. **Disclosures**. GNN reports grants, personal fees and non-financial support from Renalytix AI; non-financial support from Pensieve Health, personal fees from AstraZeneca, personal fees from BioVie, personal fees from Siemens Healthineers, personal fees from Reata, personal fees from GLG Consulting, from outside the submitted work. The remaining authors have nothing to disclose.

## Abstract

Kidney disease affects 50% of all diabetic patients; however, prediction of disease progression has been challenging due to inherent disease heterogeneity. We use deep learning to identify novel genetic signatures prognostically associated with outcomes. Using autoencoders and unsupervised clustering of electronic health record data on 1,372 diabetic kidney disease patients, we establish two clusters with differential prevalence of end-stage kidney disease. Exome-wide associations identify a novel variant in *ARHGEF18,* a Rho guanine exchange factor specifically expressed in glomeruli. Overexpression of *ARHGEF18* in human podocytes leads to impairments in focal adhesion architecture, cytoskeletal dynamics, cellular motility, and RhoA/Rac1 activation. Mutant GEF18 is resistant to ubiquitin mediated degradation leading to pathologically increased protein levels. Our findings uncover the first known disease-causing genetic variant that affects protein stability of a cytoskeletal regulator through impaired degradation, a potentially novel class of expression quantitative trait loci that can be therapeutically targeted.

## Introduction

Kidney disease is one of the most important complications associated with diabetes. Diabetic kidney disease (DKD), defined as microalbuminuria (urine albumin to creatinine ratio ≥30 mg/g) and decreased glomerular filtration rate (<60 mL/min/1.73 m^2^), affects approximately half of patients with type 2 diabetes^1^. Advancements in management of type 2 diabetes have led to a decrease in the proportion of individuals who develop DKD over the past three decades. However, due to higher disease incidence^2^, the burden of DKD is still increasing. Thus, the identification of novel kidney-specific drug targets remains critical.

The pathophysiology of DKD is multifaceted with several key signaling pathways implicated in its progression^3, 4^. It is well known that starting with the early stages of the disease, maladaptive structural changes impact glomerular function and podocyte health. Cytoskeletal remodeling, starting with cellular de-differentiation and abnormal adhesivity of podocytes^5–7^, leads to foot process effacement and eventual podocyte loss. Clinically, reduced number of podocytes is associated with progression of DKD in diabetic patients^8, 9^; however, the genomic basis of disease progression is poorly understood.

Despite the recent successes of major consortia on identifying new genetic associations with DKD^10–12^ very few cytoskeletal variants have been identified as protection or susceptibility genes against disease progression^13^. Lack of clear genetic guidance coupled with the inherent difficulty of subphenotyping the complex DKD cohort has made it difficult to develop novel precision therapeutics that target cytoskeletal remodeling. Recently, deep learning techniques have been applied to clinical and genomics datasets to uncover subphenotypes of Parkinson’s disease^14^, cancer^15, 16^, autism spectrum disorder^17^, acute kidney injury^18^, and type 2 diabetes^19, 20^. However, clustering of high dimensional electronic health record (EHR) data remains challenging due to the relative uniformity of distance measures between points in high dimensional spaces. To circumvent this “curve of dimensionality”, feature transformation techniques are often applied to decrease the number of collinear features. One such technique, autoencoders, are an unsupervised artificial neural network that compresses an input data matrix into a smaller dimension and then reconstruct the input layer^21^. The middle layer of this neural network is a representation of the input layer in a low dimensional feature space that may be used for downstream clustering algorithms.

Here, we train an autoencoder on a high dimensional dataset of DKD patients from an academic medical center with linked multimodal EHR and exome sequencing data. Using the low dimensional hidden layer from this autoencoder, we perform unsupervised clustering with a mixture of Gaussians model that accounts for population stratification^22^. Identifying the specific genetic variants associated with each resulting cluster, we find the variant rs117824875 in the *ARHGEF18* gene at exome wide significance. *ARHGEF18*, which encodes the 114 kDa protein Rho-specific guanine nucleotide exchange factor 18 (GEF18) or p114-RhoGEF, is one of the guanine nucleotide exchange factors responsible for the activation of Rho-GTPases, RhoA and Rac1^23, 24^. It was also recently reported that FERM domain proteins, EPB41L5 and EPB41L4B may be interacting with or activating GEF18 at the focal adhesion (FA) complexes^24, 25^. Single-cell RNA sequence analysis shows that *ARHGEF18* expression is specific to podocytes in the glomerulus^6^, while NephroSeq database shows that its expression correlates with proteinuria in chronic kidney disease (CKD) patients. In this study, we explore the mechanistic role of *ARHGEF18* genetic variation in podocyte morphology and function using a combination of integrated quantitative physiological assays.

## Results

### Clinical clustering identifies distinct DKD subtypes

Clinical characteristics of the patients included are presented in **Table 1**. We used mean vitals, mean laboratory values, and human phenotype ontology (HPO) terms extracted from clinical notes to identify DKD patient clusters using an unsupervised clustering strategy. DKD cases and controls were defined using phenotyping algorithms (**Supplementary Note 1, 2**). Applying a recently published clustering algorithm^22^, we adjusted for population stratification directly and identified two clusters (**Figure 1A, Supplementary Figure 1**). The silhouette score was maximal for k= 2 when applying K-means clustering (**Figure 1B**). The clusters were labelled as M (mild) and S (severe). Cluster S included 390 individuals and cluster M included the remaining 972 individuals. As compared to cluster M, cluster S was characterized by significantly greater prevalence of end stage renal disease (ESRD) (15.6% vs. 5.1%; p <0.001), slightly higher baseline serum creatinine (1.2 vs. 1.1 mg/dL; p<0.001), greater proteinuria (urinary albumin to creatinine ratio, UACR: 25 vs 15 mg/g, **Figure 1C**), and lower BMI (30.0 vs. 31.8; p=0.001) (**Table 1)**. Comparing laboratory values, cluster S had a significantly higher mean blood urea nitrogen and red distribution width and lower mean corpuscular hemoglobin concentration, plasma calcium, plasma sodium, hemoglobin, and low-density lipoprotein (**Supplementary Table 1**; FDR<0.05). Comparing HPO terms, cluster S had a greater frequency of terms related to hypervolemia, renal insufficiency, tricuspid regurgitation, vascular calcification, and pleural effusion and lower frequency of terms related to obesity, rhinitis, lower back pain, neck pain, and urinary incontinence. (**Supplementary Table 2**; FDR <0.001). Comparing vitals, cluster S had a significantly higher mean respiratory rate (18.4 vs 17.9; p<0.001) and lower mean weight (175 vs. 187 lbs.; p <0.001) (**Supplementary Table 3**).

**Figure 1:**
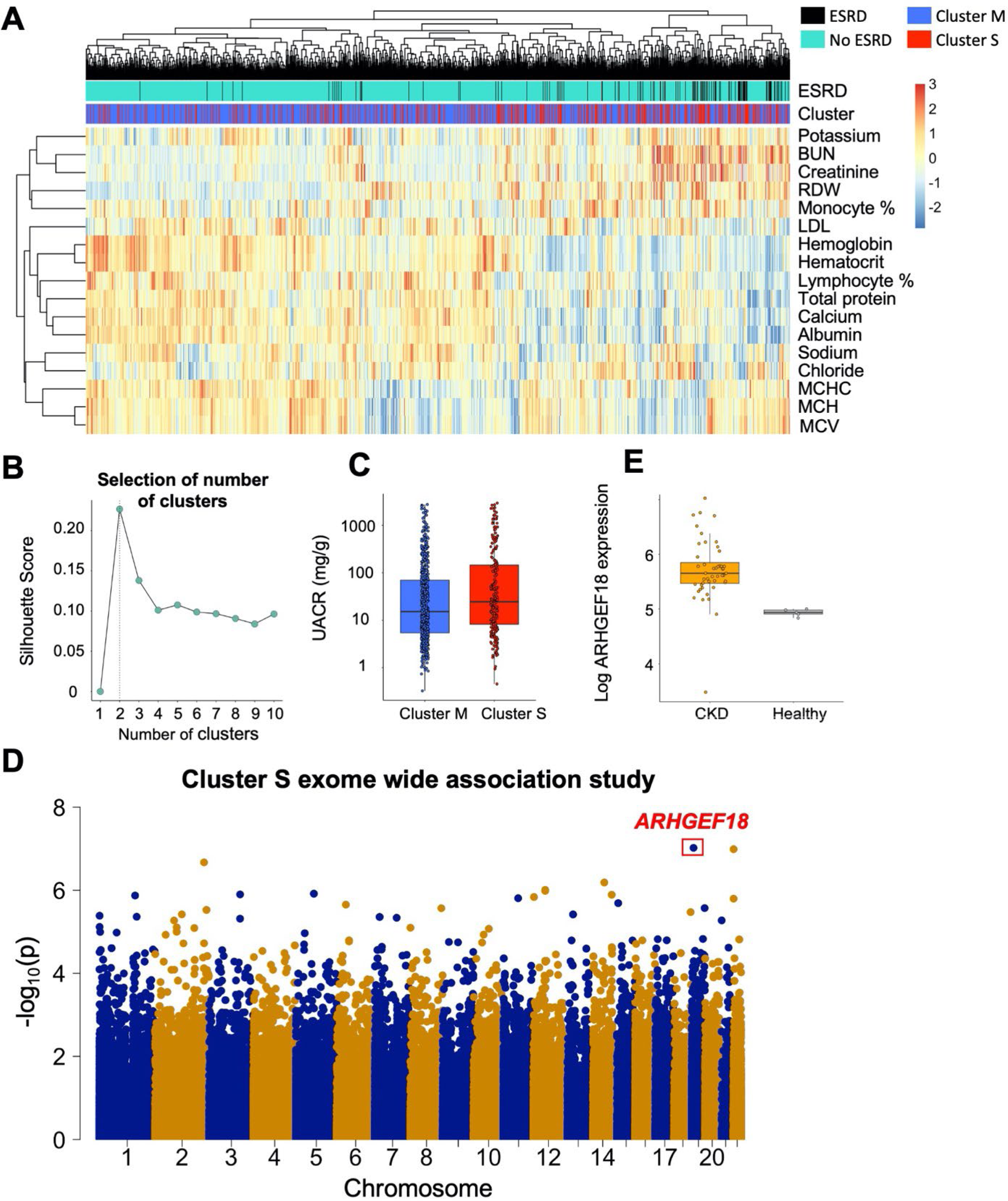
Unsupervised clustering reveals distinct DKD clinical subtypes with underlying human genetic associations. **(A)** Heatmap of laboratory values that differed significantly between clinical cluster (FDR <0.05). **(B)** Silhouette scores for clusters were computed for different values of k, the number of clusters. The value of k that maximized the silhouette score was chosen to identify the optimal number of clusters. **(C)** Baseline urinary albumin to creatinine ratio (UACR) is plotted for individuals in each cluster in BioMe. **(D)** Manhattan plot for exome wide association study comparing cluster S with DKD controls. **(E)** *ARHGEF18* mRNA expression from renal biopsy specimens is plotted for all individuals in a publicly available biopsy cohort that included patients with CKD and healthy controls.

**Table 1:** Baseline Characteristics of DKD Cohort.

|  | Cluster |  |  |
| --- | --- | --- | --- |
|  | Cluster M (N= 972) | Cluster S (N= 390) | P Value |
| Mean Age (SD) | 73.9 (9.7) | 74.5 (10.3) | 0.3 |
| Mean BMI (SD) | 31.8 (0.4) | 30.0 (0.5) | 0.001 |
| Race, n(%) |  |  | 0.04 |
| African-American | 257 (26.4) | 116 (29.7) |  |
| Asian | 15 (1.5) | 6 (1.5) |  |
| Caucasian | 105 (10.8) | 60 (15.4) |  |
| Hispanic | 536 (55.1) | 182 (46.7) |  |
| Other | 59 (6.1) | 26 (6.7) |  |
| Diabetes Clinical History |  |  |  |
| ESRD (%) | 50 (5.1) | 61 (15.6) | <0.001 |
| Renal Transplant (%) | 13 (1.3) | 11 (2.8) | 0.07 |
| Mean creatinine at enrollment (SD) mg/dL | 1.1 (0.4) | 1.2 (0.5) | <0.001 |
| Mean urinary albumin to creatinine ratio at enrollment (SD) mg/g | 175 (617) | 367 (964) | 0.28 |
| Mean age at DKD diagnosis (SD) years | 64 (10) | 65 (10) | 0.04 |
| Insulin dependent diabetes (%) | 745 (76) | 325 (83) | 0.01 |
| Medical Comorbidities, n (%) |  |  |  |
| Hypertension | 915 (94) | 354 (91) | 0.03 |
| Coronary Artery Disease | 456 (52) | 202 (52) | 0.11 |

### GEF18 dysfunction leads to kidney disease in humans

To infer a genetic basis for the clinical differences observed between clusters, we then conducted an exome wide association study. Exome sequencing was performed on peripheral blood from all individuals as previously described^26^. We compared individuals in each DKD cluster with controls from our cohort defined as individuals without a diagnosis of diabetic kidney disease. Comparing cluster M with controls, we found only two variants that met a nominal p-value threshold for significance of 5x10^-6^ (**Supplementary Table 4**). However, when we compared cluster S with DKD controls, we found 26 associated with case/control status (P <5x10^-6^; **Supplementary Table 5**, **Figure 1D**). The most significant association was for rs117824875 at exome wide significance (OR = 7.7; p = 9.56x10^-8^), a nonsynonymous variant located in *ARHGEF18*, a guanine nucleotide exchange factor. Eight (2.1%) individuals in cluster S and 191 (0.07%) DKD controls harbored a homozygous mutation (A/A) at this locus.

We validated the association of rs117824875 with DKD in the UK Biobank, a large population level biobank. Using diagnostic codes, we identified 14,660 individuals with type 2 diabetes subjected to whole exome sequencing of peripheral blood samples. Of these individuals, 187 (1.3%) were heterozygous (G/A) at the rs117824875 locus. No individuals carrying a homozygous (A/A) rs117824875 genotype were identified.

We found a significantly increased risk of DKD in rs117824875 carriers as compared to noncarriers (OR = 2.4, p = 0.04) adjusted for age, sex, and principal components genetic (**Supplementary Table 6**). Carriers also had a greater enrollment serum creatinine (beta = 0.08 μmol/L 95% CI: 0.01 – 0.16, p = 0.03) adjusted for age, sex, and five genetic principal components. Carriers also had a significantly greater enrollment urinary albumin to creatinine ratio (beta = 129 μg/mg, 95% CI: 34 – 226, p = 0.008).

To investigate the associations of *ARHGEF18* expression levels with human kidney disease, we evaluated the subsegment-specific gene expression changes associated with CKD in one of the publicly available NephroSeq renal biopsy cohorts^27^. Compared to healthy controls, patients with CKD had significantly greater whole kidney expression of *ARHGEF18* in biopsy specimens (**Figure 1E**, p < 0.001).

### Rs117824875 disrupts cytoskeletal dynamics and FA architecture

To investigate the mechanistic role of rs117824875 on podocyte function, we generated immortalized human podocyte cell lines that overexpressed either wild type (GEF18^WT^) or mutant GEF18 (GEF18^MT^) or the tag-only (EGFP) empty vector control (**Supplementary Figure 2**). We first noted that the number of GEF18^MT^ overexpression cells were significantly lower 90 minutes after replating cells cultured at 33°C. (**Figure 2A**). This was independent from the tissue culture substrate (**Figure 2B**), which suggested an adhesive abnormality not linked to a specific extracellular ligand receptor. While we did not observe an adhesion abnormality in the GEF18^WT^ cells, cell and nuclear area were significantly decreased as a result of *ARHGEF18* overexpression in GEF18^WT^ and GEF18^MT^ cells. (**Figure 2C**). This effect was also more pronounced in GEF18^MT^ cells as compared to GEF18^WT^ cells. In terminally differentiated cells 90 minutes after replating, we noted that GEF18^MT^ cells consistently showed significantly decreased adhesion compared to the GEF18^WT^ and EGFP groups (**Supplementary Figure 3A**).

**Figure 2:**
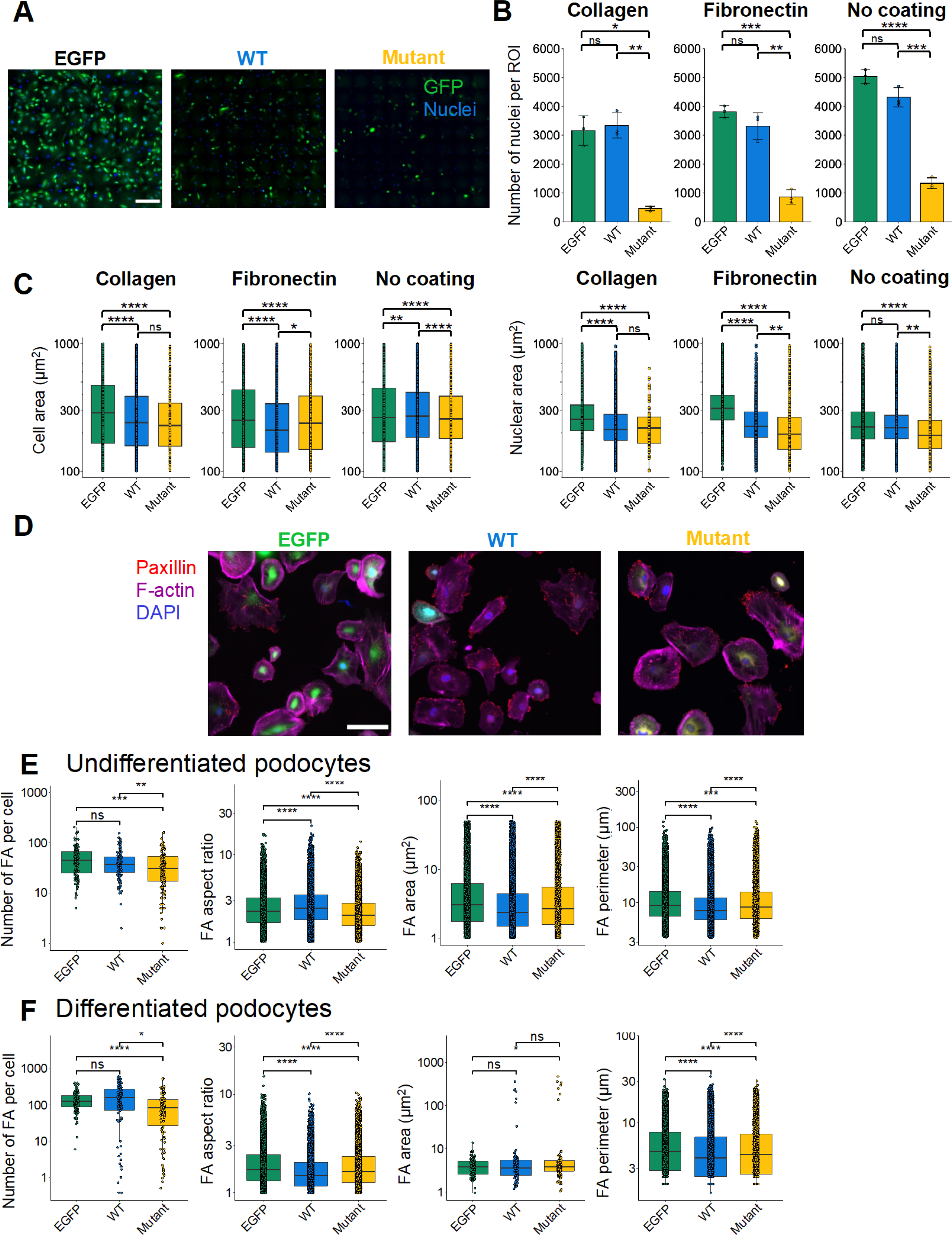
Rs117824875 impairs podocyte focal adhesions and morphology. Immortalized proliferating human podocyte cell lines that overexpress either **(A)** GEF18^WT^, GEF18^MT^, or a control EGFP vector were plated on collagen, fibronectin, or uncoated plastic dishes. GEF18^MT^ cells display **(B)** decreased cellular adhesion 90 minutes after replating, as well as **(C)** decreased cellular and nuclear area. **(D)** Representative immunofluorescence images of focal adhesions in terminally differentiated podocytes plated on collagen coated dishes and imaged using Leica DMi8 Infinity TIRF microscope with a 63X TIRF objective. **(E)** Focal adhesion morphometrics from proliferating cells measured after two days in culture at 33°C. **(F)** Focal adhesion morphometrics from differentiated cells measured after 10 days of differentiation at 37°C. All experimental groups were taken from a minimum of three plates. Significance was evaluated using a Kruskal-Wallis test followed by a post hoc Tukey test (****p<0.0001, ***p<0.001, **p<0.01, *p<0.05).

However, we did not see a consistent trend in the cell and nuclear area of the terminally differentiated cells (**Supplementary Figure 3B, C**). Using previously established high-content analytics^28^ on total internal reflectance fluorescence (TIRF) microscopy images (**Figure 2D**), we quantified focal adhesion-related morphological properties in podocytes cultured on collagen coated coverslips for two days at 33°C or differentiated for ten days at 37°C. In the proliferative context under 33°C, GEF18^MT^ cells displayed significantly fewer FA complexes per cell, which were also shorter as compared to control EGFP cells and GEF18^WT^ cells (**Figure 2E, Supplementary Figure 4A**). In terminally differentiated cells, we observed similarly decreased total FAs in GEF18^MT^ cells as compared to GEF18^WT^ (**Figure 2F, Supplementary Figure 4B**).

Given the aberrant morphometrics observed in GEF18^MT^ cells, we investigated the effect of *ARHGEF18* overexpression on activation of Rac1 and RhoA, small GTPases with canonical roles in cytoskeleton structure and function^29, 30^. *ARHGEF18* overexpression led to increased Rac1 and RhoA activity, with a more prominent effect in GEF18^MT^ cells as compared to GEF18^WT^ cells (**Figure 3A**). Consequently, in proliferating cells imaged after 48 hours in culture we observed a small but significant reduction in mean stress fiber length in GEF18^MT^ and GEF18^WT^ cells as compared to EGFP control (**Figure 3B, C**). There was also a small but significant reduction in the number of fibers in the GEF18^WT^ cells, which did not translate into a reduction in cytoskeletal coverage (**Figure 3C, D**). We also note that cell and nuclear morphometrics obtained from podocytes cultured for 48 hours showed no significant difference between the GEF18^MT^ and EGFP (**Supplementary Figure 5**). While this may seem contradictory with the aberrant spreading phenotype observed immediately after replating (**Figure 2C**), it is likely due to survivorship bias, where cells overexpressing *ARHGEF18* at high levels, particularly the GEF18^MT^ cells, are less likely to survive. Indeed, viability assay performed during the late spreading phase shows significantly increased cell death in GEF18^MT^ podocytes in a dose dependent manner (**Supplementary Figure 6**).

**Figure 3:**
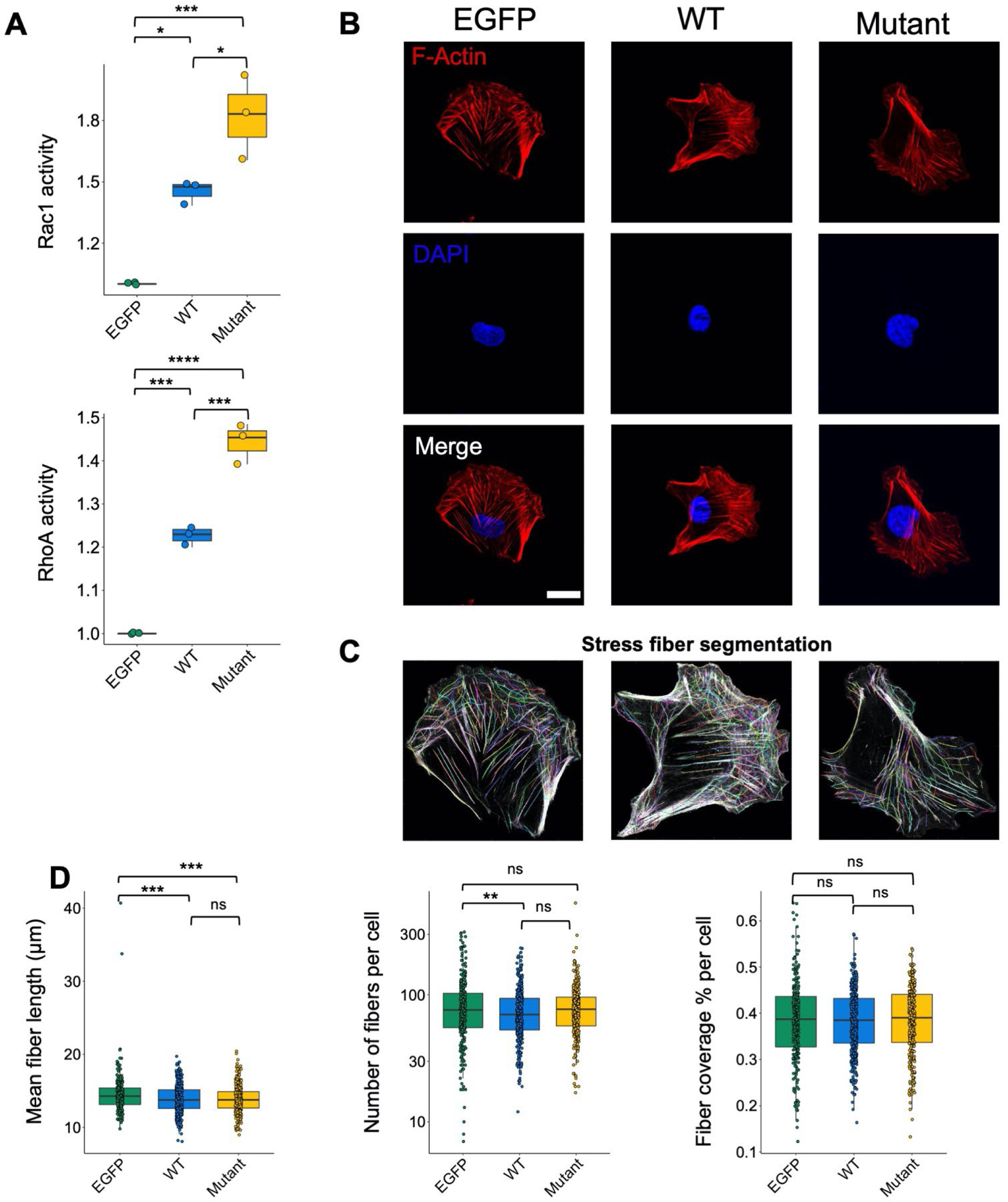
Effect of rs117824875 on podocyte cytoskeletal organization. **(A)** Normalized Rac1 and RhoA activation measured by G-LISA. **(B)** Representative images of actin stress fibers obtained using Zeiss LSM880 Airyscan laser scanning confocal microscope with a 1.4NA 63X oil objective. **(C)** Representative stress fiber segmentation image as generated by Cellpose. **(D)** Stress fiber properties were quantified using a previously published image processing pipeline43. Significance was evaluated using a Kruskal-Wallis test followed by a post hoc Tukey test (****p<0.0001, ***p<0.001, **p<0.01, *p<0.05).

### Rs117824875 alters subcellular distribution and dynamics of GEF18

Given the significant impact of GEF18 in FA architecture, we used live TIRF microscopy to determine if spatiotemporal subcellular localization of GEF18 was altered in cell lines expressing either WT or mutant protein fused with an mClover tag. There were no major phenotypic differences in cell lines overexpressing fluorescent fusion version of GEF18 compared to the unlabeled protein. However, live-cell TIRF imaging of proliferating cells over a period of six hours showed that in GEF18^MT^ cells, GEF18 was highly localized to the periphery as compared to largely perinuclear localization in GEF18^WT^ cells (**Figure 4A, B, Supplementary videos 1-4**). In terminally differentiated cells, we observed a similar peripheral localization of GEF18 in GEF18^MT^ cells as compared to GEF18^WT^ cells during live TIRF imaging, however, the effect was not as significant as seen in the proliferative context (**Supplementary Figure 7A, Supplementary videos 5-8**). We note that this difference between GEF18^MT^ and GEF18^WT^ cells was not readily observable in fixed cells suggesting that rs117824875 mostly impacts GEF18 protein localization in the context of FA dynamics and turnover (**Figure 4C**, **Supplementary Figure 7B**).

**Figure 4:**
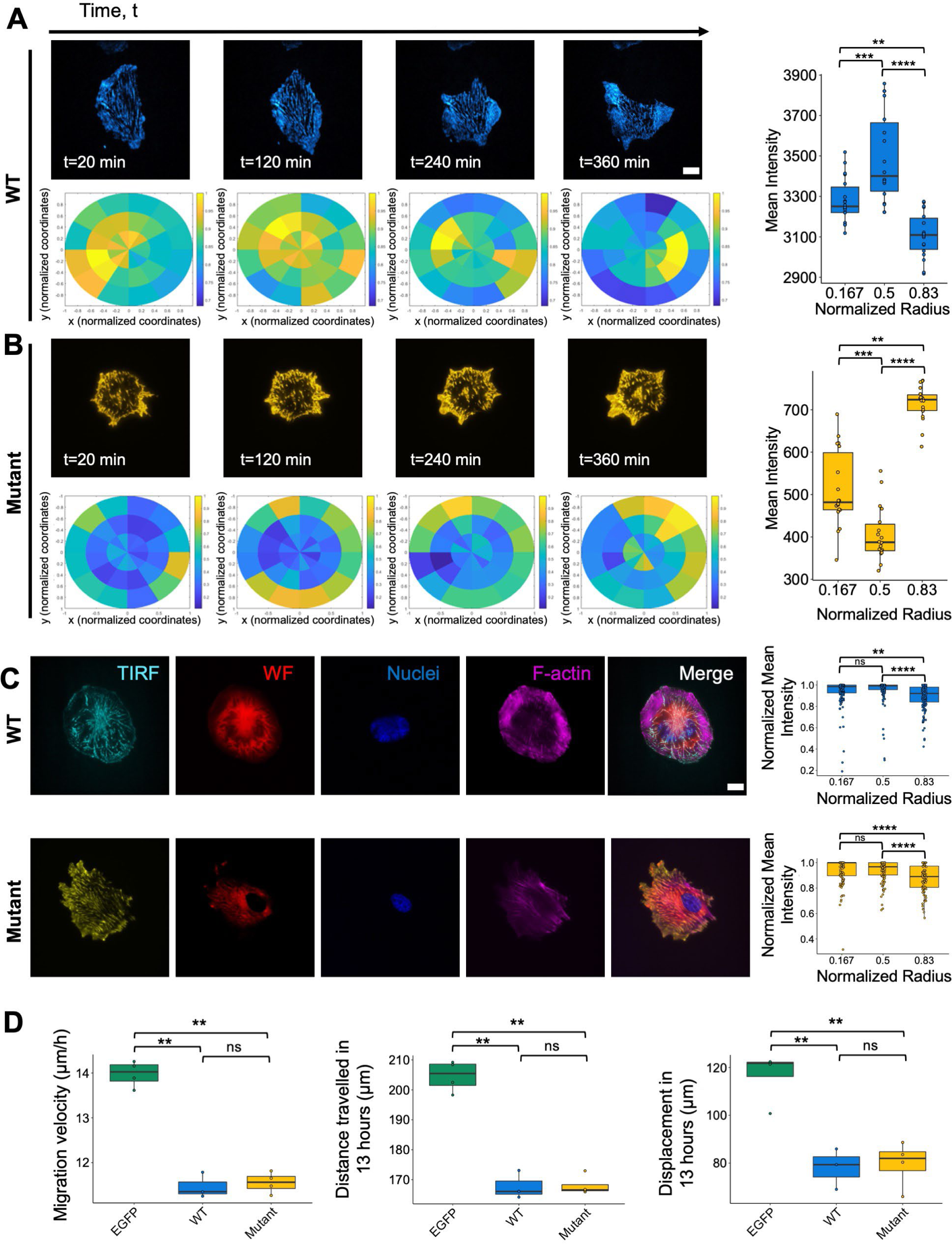
GEF18 subcellular localization and cellular motility are altered by rs117824875. Representative timelapse TIRF images in **(A)** GEF18^MT^ and **(B)** GEF18^WT^ mClover-fusion cells under the proliferative context. GEF18 protein subcellular localization was quantified by measuring mean pixel intensity as a function of radial distance of the center of each cell averaged over the timelapse. **(C)** Representative immunofluorescence staining of fixed GEF18^WT^ and GEF18^MT^ stained for nuclei (blue) and F-actin (purple). GEF18 subcellular localization was quantified using normalized mean intensities for each radial bin. **(D)** GEF18^MT^ and GEF18^WT^ cells display decreased migration velocity, distance travelled, and net displacement measured by tracking the centroids of cells in live-cell imaging analyses. Significance was evaluated using a Kruskal-Wallis test followed by a post hoc Tukey test (****p<0.0001, ***p<0.001, **p<0.01, *p<0.05).

Since FA dynamics were substantially altered by rs117824875, we also tracked undirected cellular motility of *ARHGEF18* overexpressing and control cells. On average, both GEF18^MT^ and GEF18^WT^ cells displayed decreased migration velocity and traversed a smaller distance during overnight culture (13 hours) as compared to GFP control cells (**Figure 4D**). We note that in terminally differentiated cells, no significant difference in migration velocity, net displacement, or distance travelled were observed (**Supplementary Figure 8**).

### Rs117824875 impacts GEF18 degradation and protein homeostasis

Several potential mechanisms may underlie the differences in cellular dynamics and GEF18 function observed in GEF18^MT^ cells. Using transfection experiments, we observed that the mutant cells displayed a higher expression of GEF18 compared to the WT cells at the same transfection dose (**Supplementary Figure 6B)**. We also noticed that GEF18^MT^ cells expressed significantly more GEF18 protein compared to GEF18^WT^ and EGFP control cell lines in both proliferating and differentiated podocytes (**Supplementary Figure 9**). We therefore investigated the effect of rs117824875 on GEF18 protein stability. We first inhibited protein synthesis by treating cells with cycloheximide (CHX) and quantified GEF18 protein levels at subsequent time points. Following CHX treatment, GEF18 protein levels were significantly reduced in both GEF18^MT^ and GEF18^WT^ cells (**Figure 5A, B**). However, GEF18^MT^ cells displayed significantly slower reduction of GEF18 levels following CHX treatment, suggesting slower degradation kinetics (**Figure 5B**). Since ubiquitination and autophagy are the major pathways for protein degradation within the cell, we then sought to determine which pathway was selectively impaired by rs117824875 (**Figure 5C**). Inhibition of degradation pathways using small molecule inhibitors showed an increased accumulation of GEF18 protein in the GEF18^MT^ cells. Using co-immunoprecipitation, we observed significantly less GEF18 ubiquitination in GEF18^MT^ cells as compared to GEF18^WT^ cells (**Figure 5D**). These data show that the GEF18 mutant protein displays increased protein stability against ubiquitin mediated degradation in GEF18^MT^ cells.

**Figure 5:**
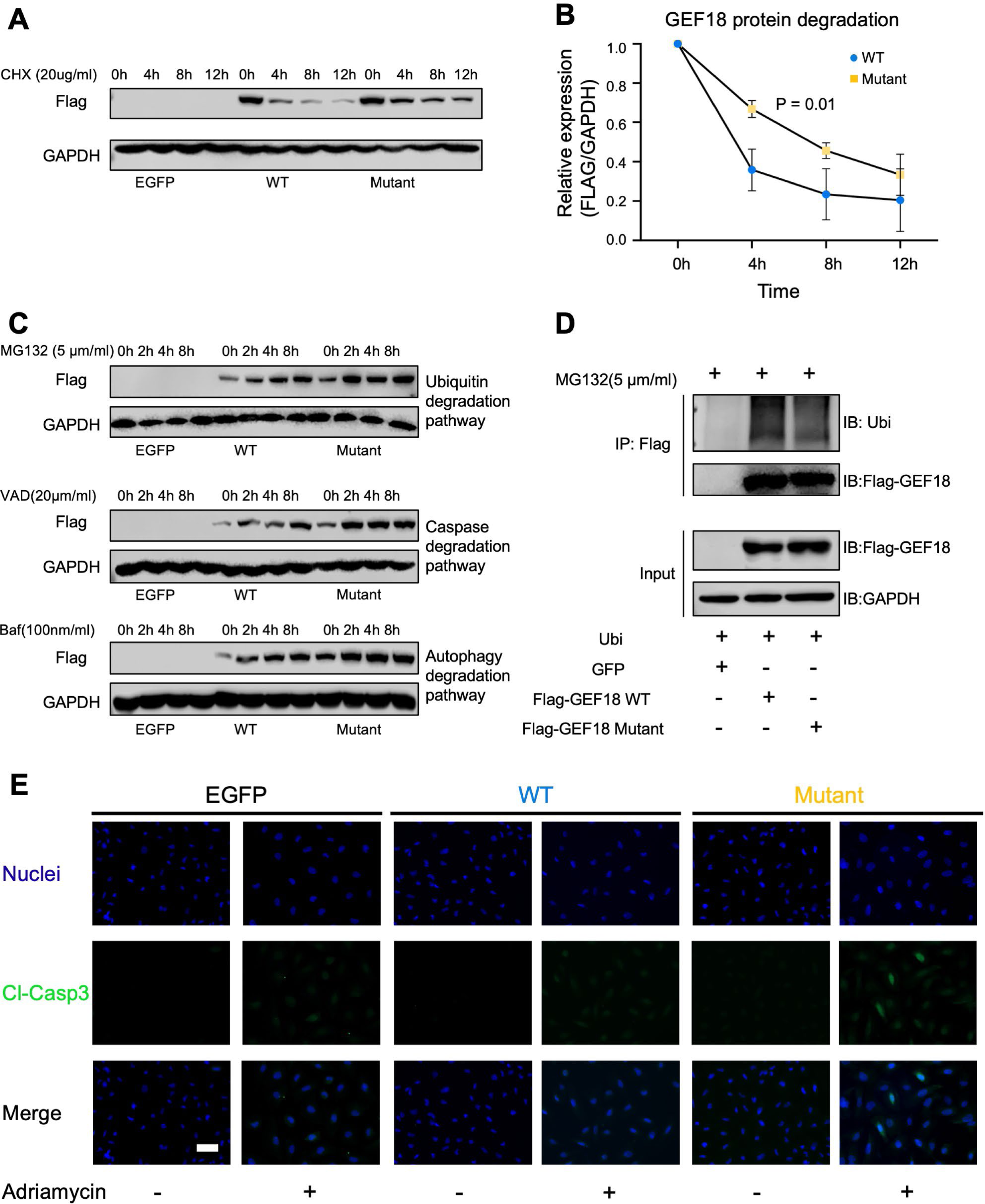
Rs117824875 enhances GEF18 protein stability and induces apoptosis. **(A)** Immortalized proliferating human podocyte cell lines that overexpress either FLAG-GEF18^WT^, FLAG-GEF18^MT^, or a control GFP vector were treated with cycloheximide. **(B)** GEF18 protein expression was quantified at several time points. Statistical significance was evaluated by fitting a linear regression model adjusted for timepoint. **(C)** Western blot of FLAG-GEF18 at different times after applying small molecule inhibitors of protein degradation pathways. **(D)** Immunoprecipitation of ectopically expressed FLAG-tagged GEF18 precipitates greater ubiquitin in GEF18^WT^ cells as compared to FLAG-GEF18^MT^ cells. **(E)** Representative images of immunofluorescence staining of nuclei (blue) and cleaved caspase-3 (green) in FLAG-GEF18^WT^ and FLAG-GEF18^MT^ cell lines.

Since aberrant cell adhesion is known to be associated with lowered podocyte resilience under disease conditions^31^, given the altered protein stability and FA dynamics in GEF18^MT^ cells, we investigated whether rs117824875 induced an increased apoptotic response in podocytes under stress conditions We observed significantly greater cleaved caspase-3 expression in GEF18^MT^ cells as compared to GEF18^WT^ cells after Adriamycin (ADR) treatment, indicative of increased propensity to undergo apoptosis (**Figure 5E**).

## Discussion

It has been recognized that genetic factors play a major role in DKD, which may explain why only a portion of diabetic patients develop DKD^32, 33^. However, large genetic association studies, such as FIND^34^ and GoKinD^10^, have failed to demonstrate any major loci associated with proteinuria and eGFR in DKD. Recently, a large Genome wide association (GWAS) study revealed a protective role of collagen IV variants for DKD^11^. Here, we demonstrate the ability of a deep learning-based clustering strategy to identify two distinct subphenotypes of DKD patients using a multiethnic high-dimensional EHR dataset. Leveraging this hidden heterogeneity, we then identify a novel cluster-specific genetic association with a gain of function exonic variant in *AHRGEF18,* a key regulator of Rac1/RhoA activity. We replicate the association of this variant with DKD and renal function in an external dataset and find increased renal expression of *ARHGEF18* in biopsy specimens of CKD patients. In an *in vitro* model system, we then demonstrate that this mutation affects cytoskeletal organization, focal adhesion dynamics, Rac1/RhoA activity, protein localization, and protein degradation. Even though this variant has a low minor allele frequency in the general population, it still represents a significant number of patients due to the large DKD population. Therefore, development of a therapeutic that targets this variant may have significant clinical utility.

The high dimensionality of EHR features makes applying conventional distance-based clustering algorithms, such as K-means, difficult. In this work, we leveraged autoencoders to reduce the dimensionality of the input feature space. By constraining the latent representation to have a smaller dimension than the input layer, we generated a minimal set of highly salient features that both preserve information and enable clustering. Given the well-recognized effect of population structure in confounding biological subtype identification^35^, we used a novel clustering method that accounts for genetic structure. By utilizing this approach, the resulting clusters likely have genuinely distinct genetic drivers and pragmatic importance.

Comparing the two clinical clusters, we found a three-fold difference in the rate of end-stage kidney disease (5% vs 15%), suggesting practical importance to our clustering strategy. Although replicating these specific clusters is difficult due to systematic differences in EHR variable distributions, future work may broadly define subtypes using aggregated data from multiple cohorts. One of the goals of our work was to demonstrate the performance of a deep learning feature selection strategy that can define subtypes with unique genetic drivers.

We also demonstrate that rs117824875 is associated with DKD diagnosis and renal function in the UK Biobank. Since clinical notes were not available in UK Biobank and the patient populations differed significantly between UK Biobank and Mount Sinai BioMe biobank, we were unable to reproduce the clusters themselves. However, a significant impact on renal function suggests that this variant has a clinical impact in multiple populations.

GEF18 is a known activator of both Rac1 and RhoA^36–38^. Since *ARHGEF18* is selectively expressed in murine podocytes and has been shown to regulate actin organization and cellular morphology *in vitro*^25, 39^, we hypothesized that the cluster S associated rs117824875 mutation may play a role in driving podocyte dysfunction. To study the functional difference between the WT and mutant protein, we generated GEF18^WT^ and GEF18^MT^ overexpressing immortalized human podocyte cell lines. Overexpression of GEF18^MT^ resulted in decreased FAs, cellular area, and nuclear area, consistent with a prominent role in regulating podocyte morphology. Decreased FAs may also be promoting podocyte apoptosis through anoikis. In GEF18^MT^ cells, there was also decreased stress fiber length but no difference in stress fiber coverage per cell. This may be a result of the shift to peripheral localization of GEF18 in GEF18^MT^ cells. We demonstrate a significant increase in activation of both RhoA and Rac1 in *ARHGEF18* overexpressing cells. This effect was stronger in GEF18^MT^ cells as compared to GEF18^WT^ cells indicating a functional consequence of the rs117824875 mutation. Using live-cell imaging, we also demonstrate impaired FA dynamics and altered cell motility in GEF18^MT^ cells, further supporting a key regulatory role for the *ARHGEF18* variant in kidney podocytes. Although we acknowledge our in vitro experiments were performed under normal glucose conditions which is not representative of a diabetic kidney environment, we observed significant differences in cell and focal adhesion morphometrics in addition to increased GTPase activation. These data show that the effect of the mutation on protein function is significantly detrimental to podocyte health even at baseline conditions.

There are several potential causes for the functional difference in GEF18 protein function induced by rs117824875; however, decreased ubiquitin mediated degradation leading to enhanced protein stability is potentially the most unique and translationally significant aspect. This mechanism potentially represents a druggable target. Highly selective degradation of target proteins by proteolysis targeting chimeras (PROTACs) has been exploited as a therapeutic strategy in several cancers^40^. PROTACs are bifunctional molecules consisting of a specific E3 ligand and a ligand that binds a protein of interest. These molecules are naturally catalytic and recruit the cell’s endogenous E3 ligase, making them effective at lower dosages with fewer adverse events^41^. Given these favorable properties, targeting GEF18 degradation by PROTAC-mediated recruitment of a podocyte specific E3 ligase may be a potential strategy to prevent RhoA/Rac1 overactivation and podocyte loss.

Recent work has identified several disease-associated genetic variants with notable effect on gene expression^42^. These expression-quantitative trait loci (eQTL) are most commonly in regulatory regions and increase disease risk by affecting mRNA levels. In this work, we find that rs117824875 is a DKD specific eQTL. Unlike other disease associated eQTLs, however, rs117824875 drives kidney disease by inducing a dysfunctional rate of GEF18 protein degradation. Since rs117824875 impairs protein stability, *ARHGEF18* overexpressing cells are likely undergoing apoptosis at a faster rate through a mechanism of natural selection. In fact, when cultured for 10 days under thermoshifted (37°C) conditions, terminally differentiated GEF18^MT^ cells have significantly lower GEF18 expression since cells with high GEF18 are more likely not to survive in culture. As such, most biochemical and functional differences observed in proliferating cells were not observed in differentiated cells through 37°C thermoshifting given the natural pruning of cells with high GEF18 expression (**Supplementary Figure 5,6**).

In conclusion, rs117824875 represents a novel rare exonic gain of function variant in the *ARHGEF18* gene that drives podocyte dysfunction through increased protein stability. Future work aimed at pharmacological inhibition of GEF18, or its increased degradation, may help regulate the balance of RhoA-Rac1 pathways, leading to improved preservation of podocyte numbers as well as function in DKD patients.

## Acknowledgements

We acknowledge funding from NIH R01DK118222 (E. U. Azeloglu). In addition, J. C. He is supported by NIH R01DK109683, R01DK122980, R01DK129467, P01DK56492, and VA Merit Award I01BX000345; G. N. Nadkarni is supported by NIH R01DK127139 and R01HL155915; G. L. Gusella is supported by NIH R01DK131047; K. Lee is supported by NIH R01DK117913 and R01DK129467; J. Fu is supported by NIH K01DK125614; J. C. Haydak is supported by NIH T32HD075735 NICHD-Interdisciplinary Training in Systems and Developmental Biology and Birth Defects; A. Mendoza is supported by NIH R01DK131047 Diversity Supplement. Confocal imaging was performed at the Microscopy CoRE at the Icahn School of Medicine at Mount Sinai. We acknowledge the Dean’s Flow Cytometry CORE at Mount Sinai for their assistance.

## Methods

### Feature Selection

We included mean values of seven vitals, 38 unique laboratory values (with at least one measurement in at least 95% of individuals), and 302 human phenotype ontology (HPO) terms present in the clinical notes extracted from the EHR.

### Study Population

We included patients from the Mount Sinai Bio*Me* Biobank. Briefly, the Bio*Me* is an electronic health record (EHR)-linked cohort with clinical notes and whole exome sequencing (N= 27,651). Enrollment of participants is predominantly through ambulatory care practices and is representative of New York City’s ethnically diverse patient population. Diabetic kidney disease (DKD) cases were defined as individuals with a history of both chronic kidney disease determined by a previously validated algorithm^44^ (**Supplementary Note 1**) and diabetes ascertained using International Classification of Disease (ICD) codes (**Supplementary Note 2**). Controls were defined as individuals without CKD or DKD.

### Exome Sequencing and Quality Control

Whole exome sequencing was performed on peripheral blood from 31,250 samples by Regeneron^26^. 8,761,478 GL-passed sites were sequenced. Samples flagged by Regeneron as being contaminated, having low coverage, or showing genotype-exome discordance were removed. Sites with Hardy-Weinberg equilibrium P<1 X 10^-6^ were also removed. Missingness (fraction of missing calls per sample, F_MISS) and depth of coverage were calculated using vcftools^45^. Mean missingness was 1.24 X 10^-3^. Nine samples with a missingness greater than 0.01 were excluded. Mean depth of coverage (mean across all sites) for all samples was 36.4x, with 45 samples having depth less than 30x, and a minimum depth of 27.0x.

212 samples were flagged as gender discordant. Of these, 17 samples were removed due to either contamination or low coverage status, four were removed due to being identified as duplicates, and 100 were able to be reconciled after resolving a labelling error. In addition, three were found to identify as transgender, and two were found to have possible sex chromosome abnormalities. The remaining 86 samples indicated to be gender discordant were removed. 158 duplicates samples were then removed. First and second-degree related individuals were excluded. Following quality control, 27,651 individuals were retained.

### Data Preprocessing

We utilized all laboratory values, vital signs, and human phenotype ontology (HPO) terms^46^ from our institutional electronic health records (EHR) to identify clusters. HPO terms were extracted from clinical notes using a natural language processing developed by Clinithink^47^. HPO terms were filtered using a previously published pipeline^48^. Each HPO term was encoded as a binary variable (1 if individual had HPO term in any clinical note, 0 otherwise). Only laboratory values with at least one measurement in greater than 95% of individuals were included. Vitals included systolic blood pressure, diastolic blood pressure, pulse oxygen, heart rate, height, weight, and respiratory rate. Missing laboratory values and vitals were imputed using a random forest based nonparametric imputation algorithm implemented in the *missForest* R package^49^. After imputation, mean values of each lab and vital measurement for everyone were included as input for an autoencoder model.

### Deriving Patient Representations using Deep Learning Autoencoder

A flowchart of the clustering process is provided in **Supplementary Figure 1B**. Following data preprocessing, we utilized autoencoders to reduce the dimensionality of the data before unsupervised clustering. An autoencoder is a neural network that takes a high dimensional vector as input, compresses, and then attempts to reconstructs the input layer. We trained a separate autoencoder on mean laboratory values, mean vitals and HPO terms as the scale and nature of each variable differed greatly. We used an autoencoder with three hidden layers with 10 nodes in each layer and a Tanh activation function. The architecture of the autoencoder is presented in **Supplementary Figure 1B.**

### Unsupervised Clustering

Identification of disease subtypes is routinely confounded by population structure that may be make the clinical heterogeneity of clusters. In order to account for population structure, we employed a recently published novel finite mixture of regression method, RGWAS^22^. This method adjusts for covariates assumed to have significant heterogeneity across the underlying disease subtypes. The matrix of vitals was merged with the hidden layers from each separate autoencoder (laboratory values and HPO terms) and then scaled to have mean 0 and variance of 1. We then performed unsupervised clustering using RGWAS adjusting 10 genetic principal components to account for population stratification. We varied the number of clusters, k, from two to six and then chose the k that maximized the cross-validated likelihood and minimized the entropy. To determine the stability of the clusters with the chosen k, we performed K-means clustering with the selected k and computed the Jaccard using a bootstrap resampling method. We also performed K-means clustering for values of k from two to six and computed the Silhouette score.

Clusters were visualized by plotting the first three components t-Distributed Stochastic Neighbor Embedding (t-SNE) components of the matrix used as input to clustering.

### Exome-wide Association Study

Exome wide association studies were performed comparing each cluster with non-DKD controls. Genetic associations were computed using plink^50^ by fitting a logistic regression model with parameters –mind.1 –geno 1 –maf 0.001 –adjust –glm firth-fallback. Variants with P < 5x10^-6^ were considered significant.

### Cell Culture

Conditionally immortalized human podocytes were a kind gift from Dr. Moin Saleem (University of Bristol, Bristol, UK). Cells were cultured in RPMI 1640 medium (Cat. No. 11875119, Gibco) containing 10% FBS (Cat. No. 26140079, Gibco), 1% insulin-transferrin-selenium-A supplement (Cat. No. 51300044, Gibco), 1% penicillin, and streptomycin (Cat. No. 10378016, Gibco) at 5% CO_2_ humidified environment in 33℃ under growth-permissive (GP) conditions or in 37°C growth-restrictive (GR) conditions. Podocytes were cultured in GP condition for two days or differentiated in GR condition for 10 days before experiments. All experiments were repeated at least three times for each indicated condition. For overexpression of *ARHGEF18* wild-type (WT) and mutant constructs, podocytes were transiently transfected using ViaFect reagent (Cat. No. E4981, Promega).

### Stable Overexpression of ARHGEF18 Wildtype and Mutant in Human Podocytes

Human *ARHGEF18* expression plasmids were constructed using the *ARHGEF18* ORF (NM_015318.3). pcDNA3.1-eGFP-ARHGEF18 wildtype and mutant constructs were verified with DNA sequencing and then cloned into a pNL4-3 plasmid containing FLAG tag. Lentivirus particles were generated by transfecting HEK293T cells with the expression plasmid along with the packaging and envelope plasmids. Podocytes were stably transduced with Flag-pNL4-3-eGFP-ARHGEF18 wildtype and mutant lentivirus, or control enhanced green fluorescent protein (EGFP) lentivirus. After 72 hours, cells were trypsinized and resuspended in PBS supplemented with 1% BSA and 2.5 mM EDTA. Cell sorting was performed with a BD FACSAria Fusion (BD Biosciences) according to the EGFP expression level. All further experiments were performed using stable expression cell lines. For Adriamycin (ADR) (Sigma) treatment, cells were incubated with medium containing 250ng/ml ADR for 24 hours. For CHX treatment, cells were incubated with medium containing 20ug/ml CHX for 0,1,2,4,6,8,12 hours. For MG132, VDA or Baf treatment, cells were incubated with medium containing 5 μmol/ml MG132(Cat. No. 474787, Millipore), 20 μm/ml VAD or 100 nm/ml Baf for 0,2,4,8 hours.

*ARHGEF18* WT and mutant expressing mClover fusion cell lines were generated by sub-cloning *ARHGEF18* WT and mutant into a VVPW lentiviral vector. Lentivirus particles were generated by transfecting HEK293T cells with the expression plasmid along with the packaging and envelope plasmids. Human immortalized podocytes were stably transduced and sorted based on mClover expression. Fusion protein expression was validated using fluorescence microscopy.

### Cell Viability

The cell viability was assessed using MultiTox-Fluor Multiplex cytotoxicity assay (Cat. No. G9201, Promega). 96-well plates containing 5000 cells per well were transiently transfected with *ARHGEF18* WT and mutant constructs using Viafect reagent (Cat. No. E4981, Promega). After 24 and 48 hours, 2X Multi-Fluor Multiplex Cytotoxicity reagent was added in equal volume (100 μl per well) to all the wells and mixed briefly on an orbital shaker. After 30 minutes of incubation at 37°C, fluorescence was measured using a plate reader (excitation ∼485 nm; Emission ∼ 520 nm).

### Cell Adhesion

Cell adhesion experiments were performed using immortalized human podocytes expressing EGFP, GEF18 wild type and mutant cultured under permissive and restrictive conditions. Briefly, cells were plated on 96-well imaging plates that were uncoated, collagen type I coated, or fibronectin coated and allowed to attach for 90 minutes. After 90 minutes, cells were fixed and stained with AlexaFluor 568 Phalloidin (Cat. No. A12380, Invitrogen) and Hoechst 33342 (Cat. No. 62249, Thermo Fisher). Cells were imaged on Leica DMi8 widefield microscope and the number of adhered cells were quantified based on the number of nuclei attached using Fiji. Cell and nuclear area were also quantified using a Fiji script.

### Live-cell Motility

Human immortalized podocytes were plated on collagen-coated Ibidi 8-chamber slide at a density of 2000 cells per well and maintained at 33°C for 2 days or differentiated for 10–14 days at 37°C before experiments. Live cell imaging was performed with a Leica DMi8 widefield microscope with a Pecon black-box environmental enclosure using a 20X Leica air objective with a rate of 3 frames per hour over a period of 13 hours. For data analysis, cell nuclei were segmented using Cellpose (v0.6) to generate masks for each frame in the time course, as previously described^51^. From here, the masks were run through a custom MATLAB (R2021b) script that links nuclei in one frame to the next, generating cell trajectories by solving a linear assignment problem with the distance between nuclei as the cost.

### Immunofluorescence Staining

Immortalized human podocytes expressing EGFP, GEF18^WT^ and GEF18^MT^ were cultured on collagen coated glass coverslips. Cells were fixed with 4% PFA in PBS at room temperature for 20 minutes then treated with 0.05% Triton-X (Cat. No. #T8787; Sigma Aldrich) permeabilization solution for 15 minutes at room temperature. The permeabilization solution was replaced with blocking buffer with BSA and donkey serum and incubated at 4°C overnight. Immunostaining was performed using the primary antibodies mouse anti-Paxillin mAb (Cat. No. #ma5–13356; Invitrogen) or rabbit cleaved-caspase 3 mAb (Cat. No. 9664, Cell Signaling). Thereafter, cells were incubated with fluorophore-linked secondary antibodies (568 anti-mouse IgG: from Thermo Fisher). After another wash, either AlexaFluor-568 phalloidin (when performing fusion cell experiments, catalog #A12380) or AlexaFluor-647 phalloidin (focal adhesion morphometrics experiments, Cat. No. A22287) and Hoechst 33342 (Cat. No. 62249; all from Thermo Fisher) were used to label F-actin and nuclei, respectively. After staining, coverslips were stored in PBS at 4°C and imaged within a week. Representative confocal images were obtained using a Zeiss LSM880 confocal microscope with a 63X oil objective at 1 Airy Unit.

### Total Internal Reflection Fluorescence Microscopy

TIRF microscopy was performed using a Leica DMi8 TIRF microscope and LASX (v.3.6). For focal adhesion morphometrics quantification, cells were plated on 25 mm glass coverslips at a density of 6,000-12,000 cells per condition. Paxillin and phalloidin stained coverslips were imaged in 1X PBS (Cat. No. # P36975; Thermo Fisher) at a penetration depth of 90 nm using a 1.4 NA Leica 63X oil TIRF objective. Focal adhesion area and count per cell was quantified using Fiji script as previously described^32^.

For TIRF experiments using GEF18 fusion cell lines, 6,000-8,000 cells were plated on 25 mm coverslips and used for experiments after 48 hours in culture at 33°C or differentiated for 10 days at 37°C. Cells were fixed and stained with phalloidin as described above. For live cell TIRF experiments, cells were imaged over 6 hours at 20-minute intervals. After imaging, binary cell masks of individual timepoints are generated using Fiji script. A custom developed MATLAB code was used quantify spatial localization of GEF18 during cell migration by segmenting cells into radial and angular components. Radial segmentation divides the cells into 3 segments: a radius of 0.1667 pixels shows nuclear localization, 0.5 – perinuclear and 0.83 – peripheral localization. Angular segmentation divides the cells into 12 segments, with 0 degree (or 3 o’clock position) pointing to the direction of cell movement at any given timepoint. Mean intensities are obtained for all the segments across for all time points and plotted as *imagesc*, *beeswarm* or polar plots. For intensity normalizations, a value of 1 was assigned to the segment with maximum mean intensity in each cell.

### Stress Fiber Analysis

Stress fiber analysis was performed as described by Zhang *et al*^43^. Briefly, cells were segmented from the raw image using CellPose^51^. For each cell, the bounding box was top hat filtered and the image was adjusted to saturate the top 1% and bottom 1% of all pixel values using Matlab. The individual cell image was run through a modified version of the pipeline from Zhang *et al.* allowing for batch processing without the use of a graphical user interface. The images are filtered to enhance stress fiber contrast, binarized and skeletonized. In our implementation, binarization was achieved using k-means clustering with k = 5, keeping the brightest three clusters as the binarized image. Stress fibers are then reconstructed from the skeletonized image. Final images of each cell were saved with the stress fiber overlay and manually inspected to ensure good segmentation quality.

### Western Blot Analysis

Human immortalized podocytes were seeded at a density of ∼50,000 cells in 100 mm culture dishes and allowed to grow for two days at 33°C or differentiate for 10 days at 37°C. After two days in culture or 10 days of differentiation, the cells were lysed were lysed using 50 µL of RIPA Lysis and Extraction Buffer (Cat. No. 89901, Thermo Fisher Scientific) containing Halt™ Protease and Phosphatase Inhibitor Cocktail (100X) (Cat. No. 78442, Thermo Fisher Scientific) at 2X concentration. Equal amounts of protein (30 µg) were separated on Bolt 4 to 12%, Bis-Tris, 1.0 mm, Mini Protein Gel, (Cat. No. NW04125BOX, Thermo Fisher Scientific) and transferred at 25V to a Supported Nitrocellulose Membrane, 0.45 µm (Cat. No. 1620094, Biorad) for 30 minutes. Lysate preparation and Western blotting were performed according to the standard protocol. Each lane contained 30 to 60 μg of total protein. The membrane was blocked under gentle agitation at room temperature for 30 minutes using 5% dry milk in TBS(1X). Membranes were incubated overnight with primary antibodies diluted in TBS-T 0.1% sodium azide at 4°C. Band density intensity for the protein of interest was normalized to either GAPDH or β-actin. The following primary antibodies were used in this study: FLAG (Cat. No. F1804, Sigma-Aldrich), cleaved-caspase 3 (Cat. No. 9664, Cell Signaling), Ubiquitin (Cat. No. 3936, Cell Signaling) and GAPDH (Cat. No. 2118, Cell Signaling).

### RhoA and Rac1 Activity

Human immortalized podocytes were serum starved for three consecutive days at 1% FBS in standard RPMI1640 medium. Dishes were washed with ice-cold PBS buffer twice and lysis was performed at 4°C. Lysates were equalized due to protein content and analyzed according to the manufacturer’s instruction (RhoA G-LISA activation assay kit, Cat. No. # BK124; Rac1 G-LISA activation assay kit, Cat. No. BK128, Cytoskeleton, USA).

### Statistical Analyses

After identification of DKD clusters, we compared features between clusters using the t-test and Chi-squared test for continuous and categorical variables, respectively. False discovery rate was computed using the Benjamini-Hochberg procedure. For comparison of high content image analytics, we used nonparametric Kruskal-Wallis analysis of variance with post hoc Tukey test. Significance was achieved at alpha 0.05.

## Data Availability

BioMe data, including clinical and genetic sequencing data is not publicly available due to patient privacy concerns and institutional policy. Transcriptomic data from CKD biopsy specimens is available on the Nephroseq website.

## Code Availability

Clustering analysis was performed using the publicly available RGWAS package. Downstream genetic association analyses were performed using the publicly available PLINK package.

