## Supplementary materials for "Deep learning on electronic medical records identifies distinct subphenotypes of diabetic kidney disease driven by genetic variations in the *Rho* pathway"

**Supplementary Note 1 CKD Definition**

Phenotyping algorithm used to define patients with chronic kidney disease associated with hypertension or diabetes.

See attached

**Supplementary Note 2 Diabetes Definition**

Phenotyping algorithm used to define type 2 diabetes.

See attached

**Supplementary Figures**

**Supplementary Figure 1. Unsupervised clustering**

**A)** Flowchart of data preprocessing, clustering, and downstream genetic association analyses. **B)** Architecture of autoencoders used for dimensionality reduction. The hidden layer from the HPO terms and laboratory value autoencoders were concatenated with vitals to form an input for unsupervised clustering.

### Supplementary Figure 1. Unsupervised clustering

**A**

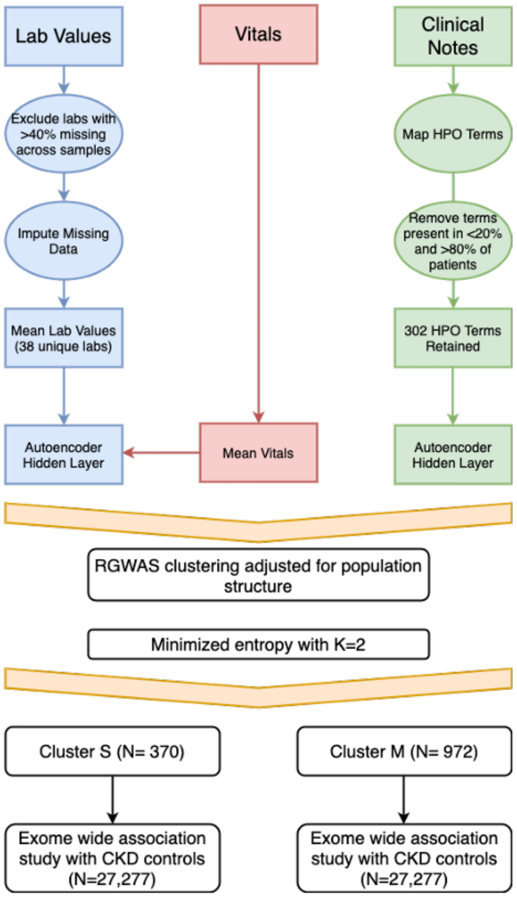


**B** **Autoencoder Architecture**

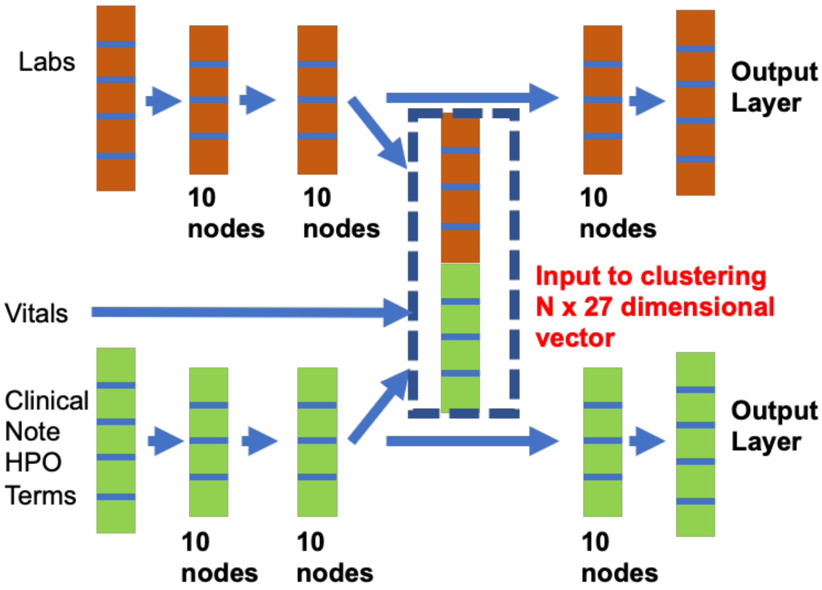

**Supplementary Figure 2. Generation of stable cell lines**
Human immortalized podocyte cells were transduced with viral vectors containing either a GFP construct, flag-GEF18$^{WT}$ construct or flag-GEF18$^{MT}$ construct. FACS was used to select cells with high GFP expression. Flag-GEF18 expression was quantified from whole cell lysates by Western blot.

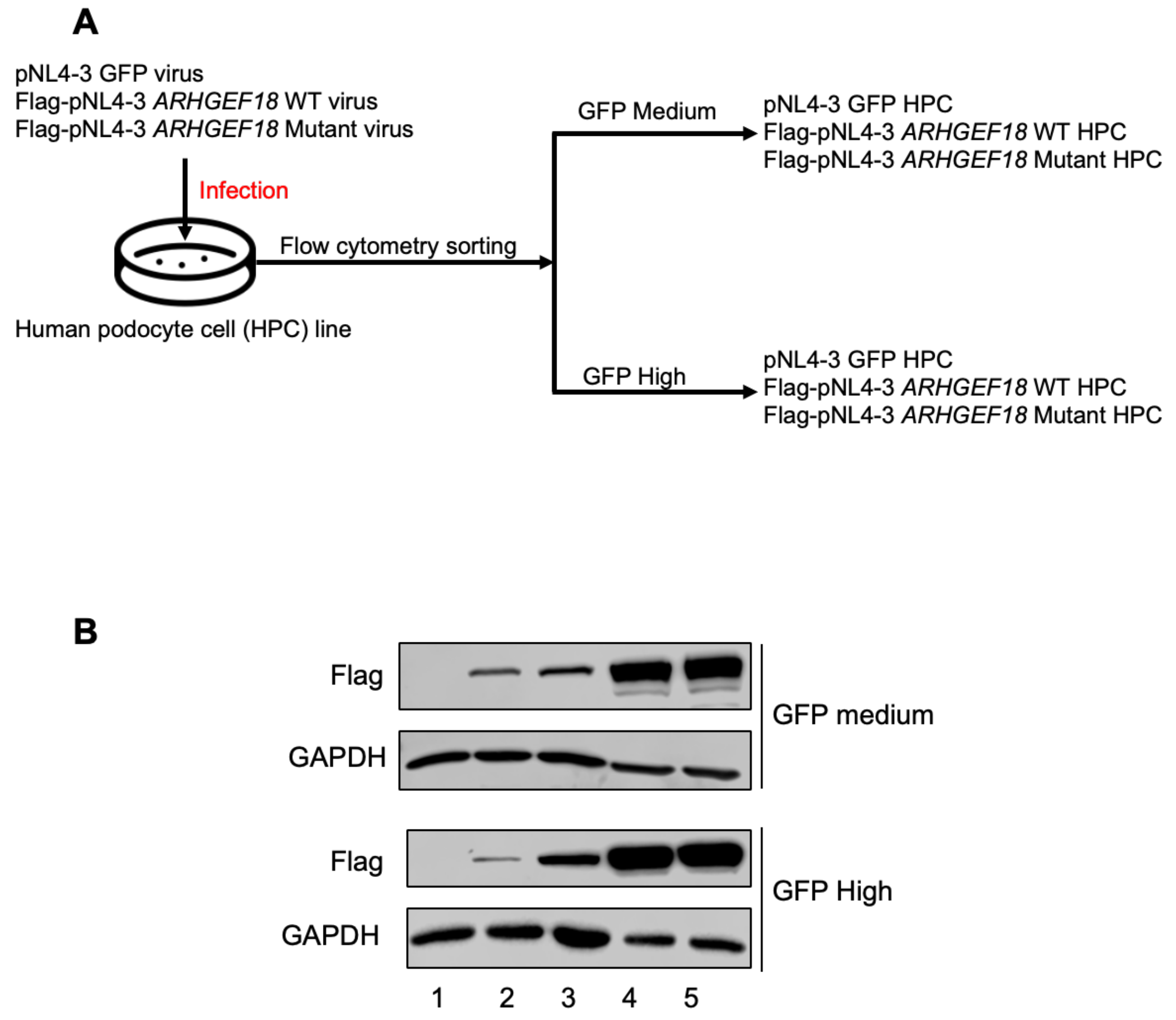


1. pNL4-3 GFP HPC stable cell line
2. Flag-pNL4-3 *ARHGEF18* WT HPC stable cell line
3. Flag-pNL4-3 *ARHGEF18* Mutant HPC stable cell line
4. HPC transfected with Flag-pNL4-3 *ARHGEF18* WT
5. HPC transfected with Flag-pNL4-3 *ARHGEF18* Mutant

**Supplementary Figure 3. Cell and nuclear morphometric analysis in terminally differentiated podocytes**

**A)** Cellular adhesion of terminally differentiated podocytes was quantified. Using a previously published image processing pipeline[1], cell and nuclear morphometrics were quantified. Significance was evaluated using a Kruskal-Wallis test followed by a post hoc Tukey test. (****p<0.0001, ***p<0.001, **p<0.01, *p<0.05).

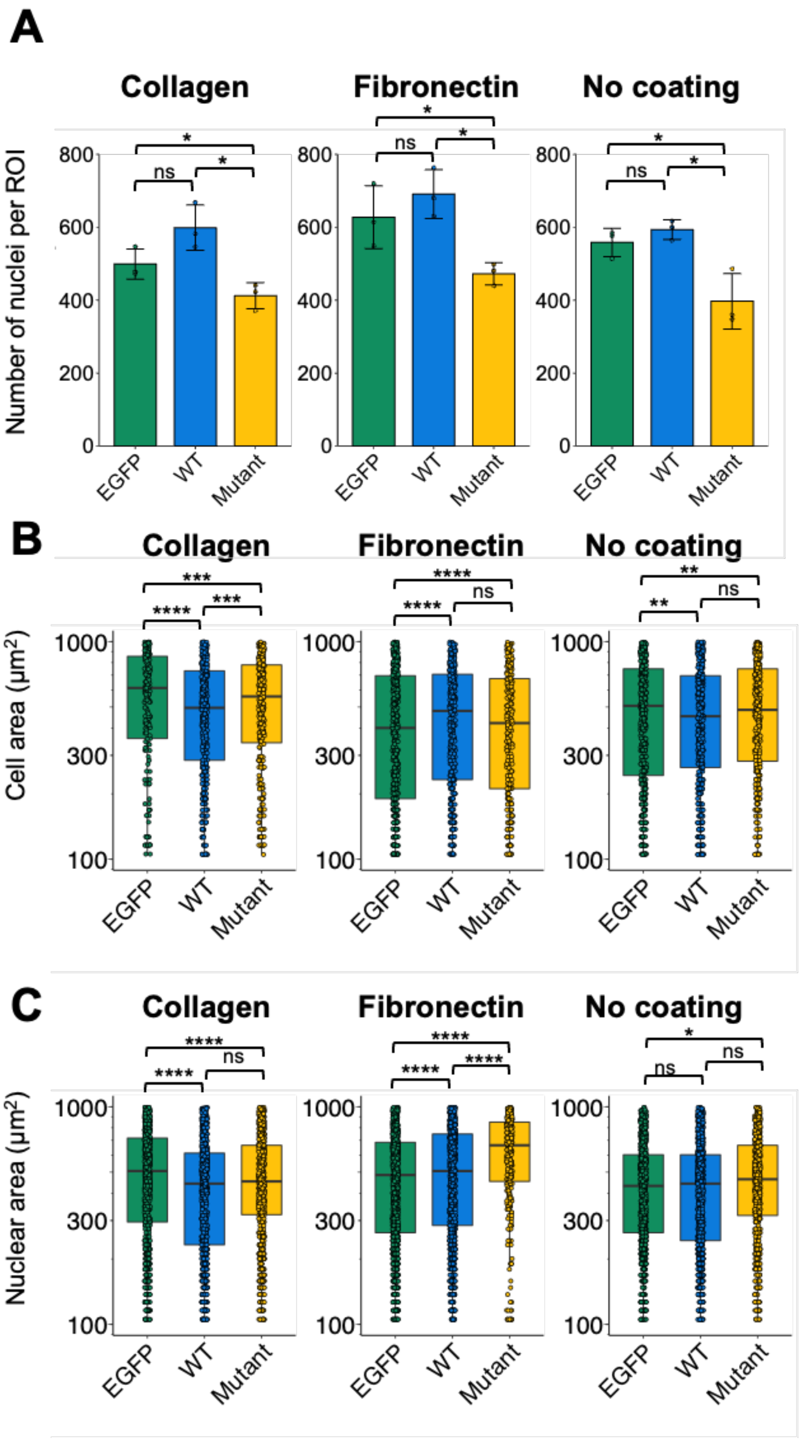

**Supplementary Figure 4. Stress fiber properties in proliferating podocytes**
**A)** Focal adhesion area per cell from proliferating cells measured after two days in culture at 33°C. **B)** Focal adhesion area per cell from differentiated cells measured after 10 days of differentiation at 37°C. All experimental groups were taken from a minimum of three plates. Significance was evaluated using a Kruskal-Wallis test followed by a post hoc Tukey test (****p<0.0001, ***p<0.001, **p<0.01, *p<0.05).

**A**

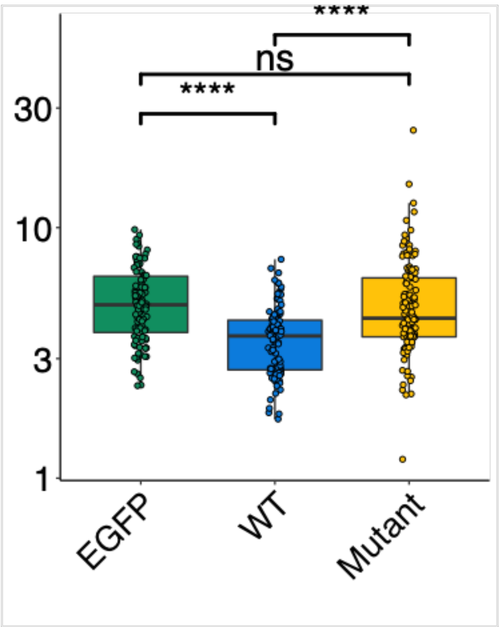


**B**

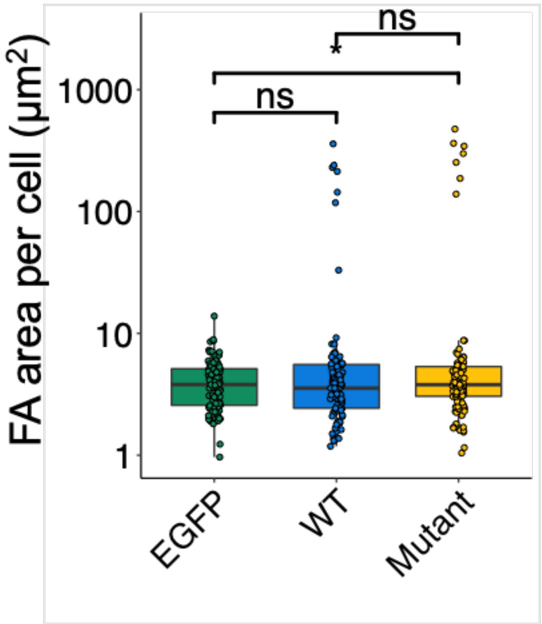

**Supplementary Figure 5. Stress fiber properties in proliferating podocytes**

Confocal microscopy was used to image stress fibers in proliferating podocytes that overexpressed either $GEF18^{WT}$, $GEF18^{MT}$, or a control GFP vector. Cell area, cell perimeter, nuclear area, and nuclear perimeter were quantified using a previously published pipeline[1]. Significance was evaluated using a Kruskal-Wallis test followed by a post hoc Tukey test (****p<0.0001, ***p<0.001, **p<0.01, *p<0.05).

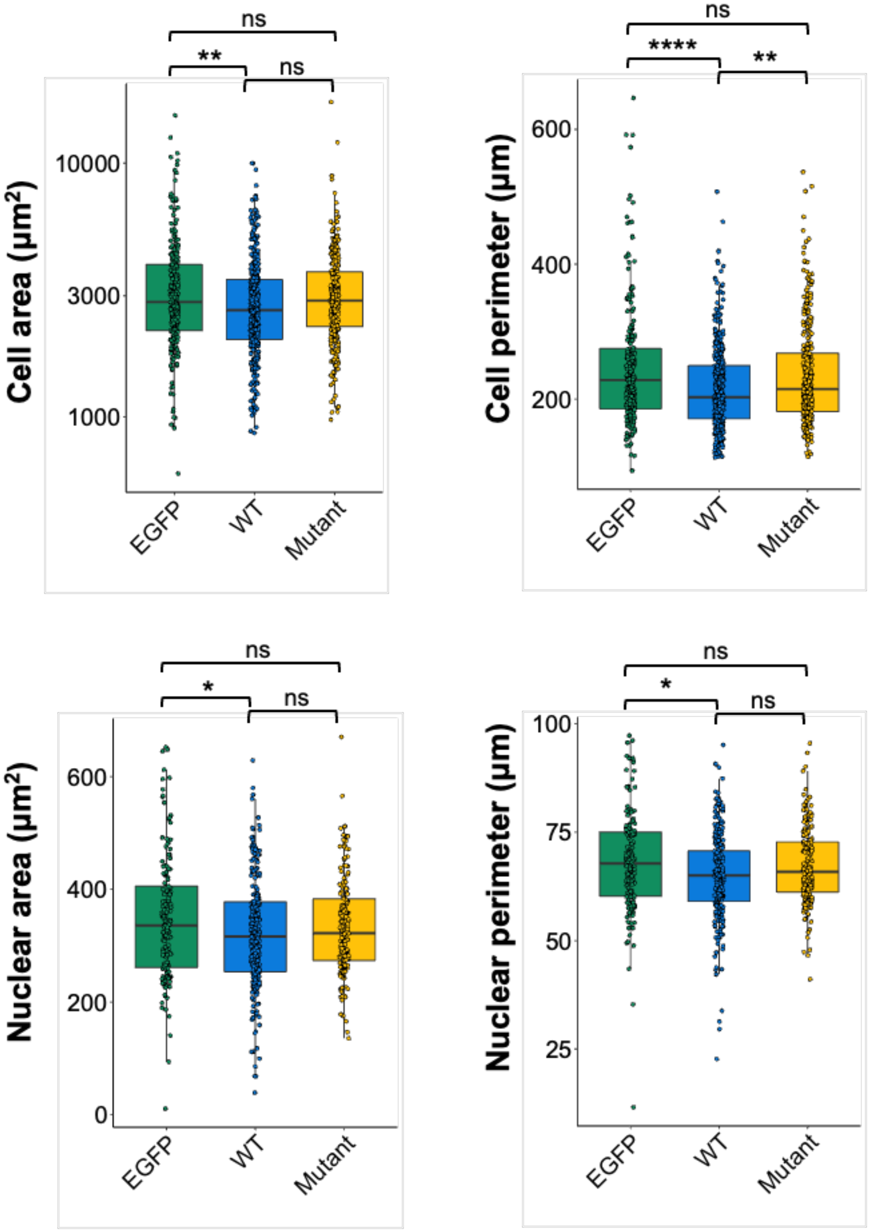

**Supplementary Figure 6. Cell viability of proliferating podocytes**

A) Viability of proliferating podocytes that overexpressed either GEF18$^{WT}$ or GEF18$^{MT}$ was measured using the MultiTox-Fluor Multiplex cytotoxicity assay at 24 and 48 hours in culture. Cell viability was computed relative to viability of podocytes transfected with a control GFP vector. B) Protein expression for cell lines transfected with different doses of plasmid.

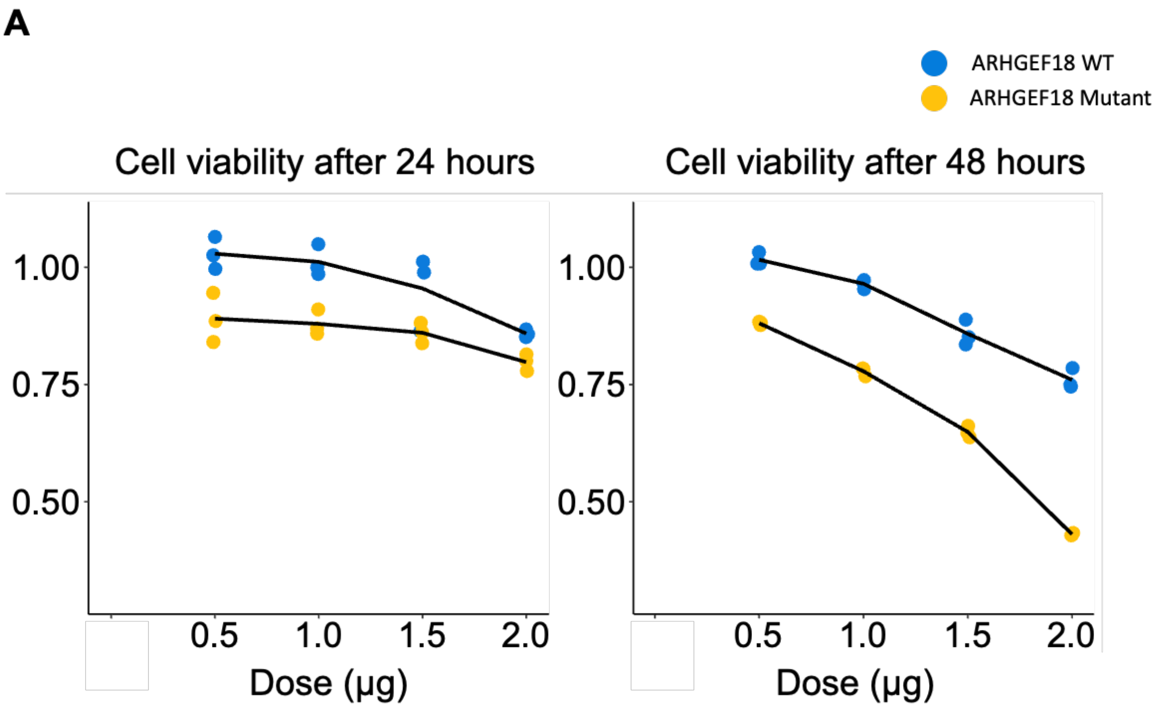


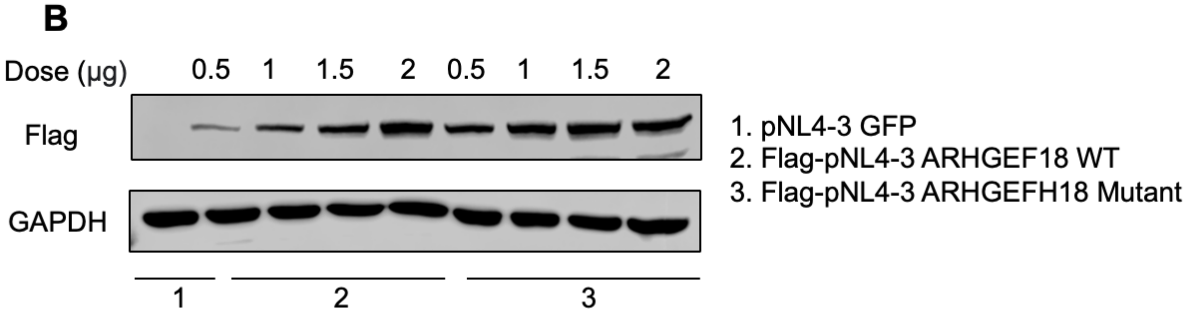

**Supplementary Figure 7. GEF18 subcellular localization in terminally differentiated podocytes**

TIRF microscopy was used to determine GEF18 subcellular localization in $GEF18^{MT}$ and $GEF18^{WT}$ mClover-fusion cells after undergoing terminal differentiation in culture. **A)** represents live cell imaging and **B)** represents fixed cells. Significance was evaluated using a Kruskal-Wallis test followed by a post hoc Tukey test (****p<0.0001, ***p<0.001, **p<0.01, *p<0.05).

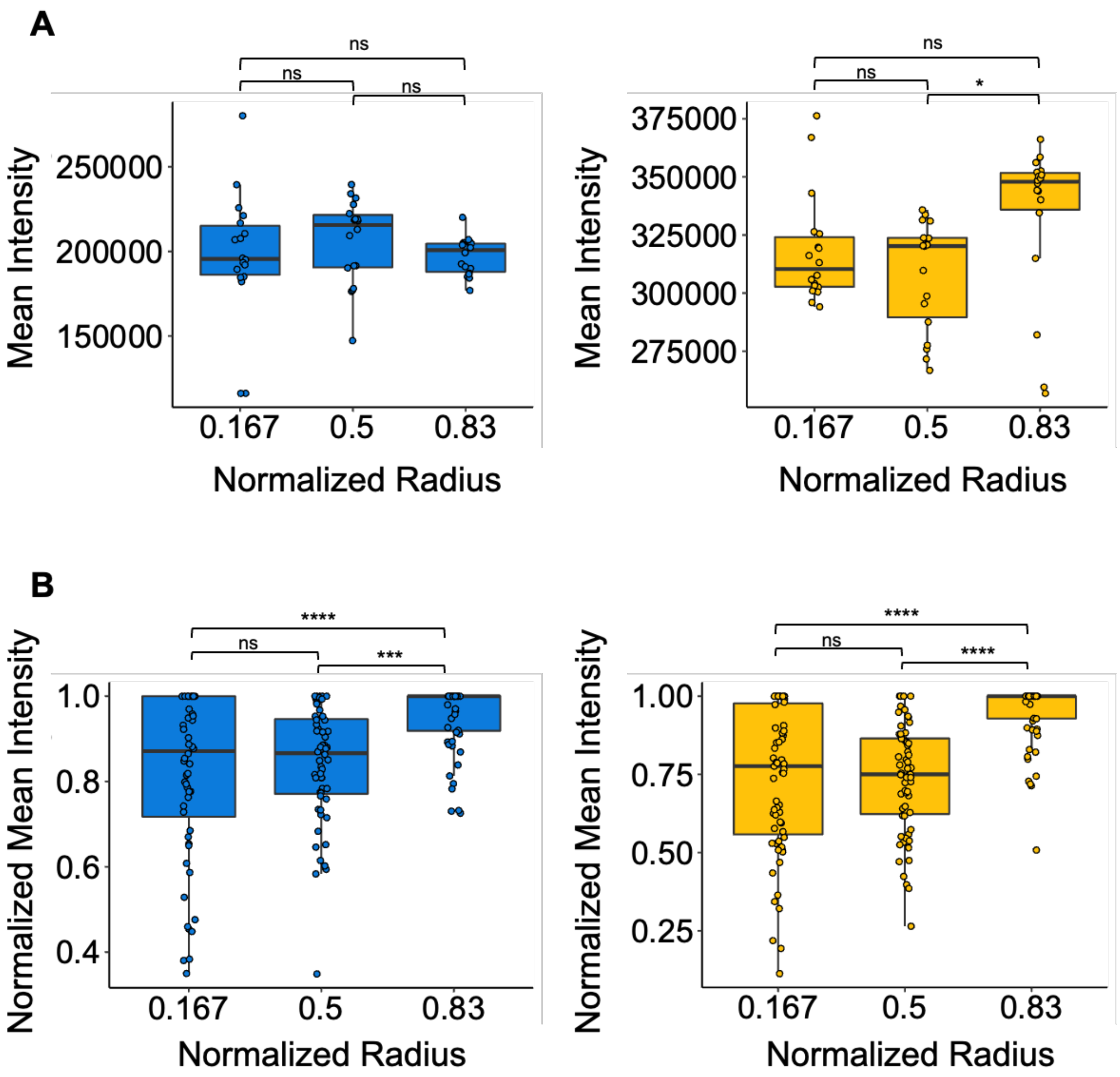

**Supplementary Figure 8. Cell migration in terminally differentiated cells**

Migration velocity, net displacement, and distance travelled was quantified from cells that overexpress either $GEF18^{WT}$, $GEF18^{MT}$, or control GFP vector that were imaged over a period of 13h using a 20X air objective. Significance was evaluated using a Kruskal-Wallis test followed by a post hoc Tukey test (****p<0.0001, ***p<0.001, **p<0.01, *p<0.05).

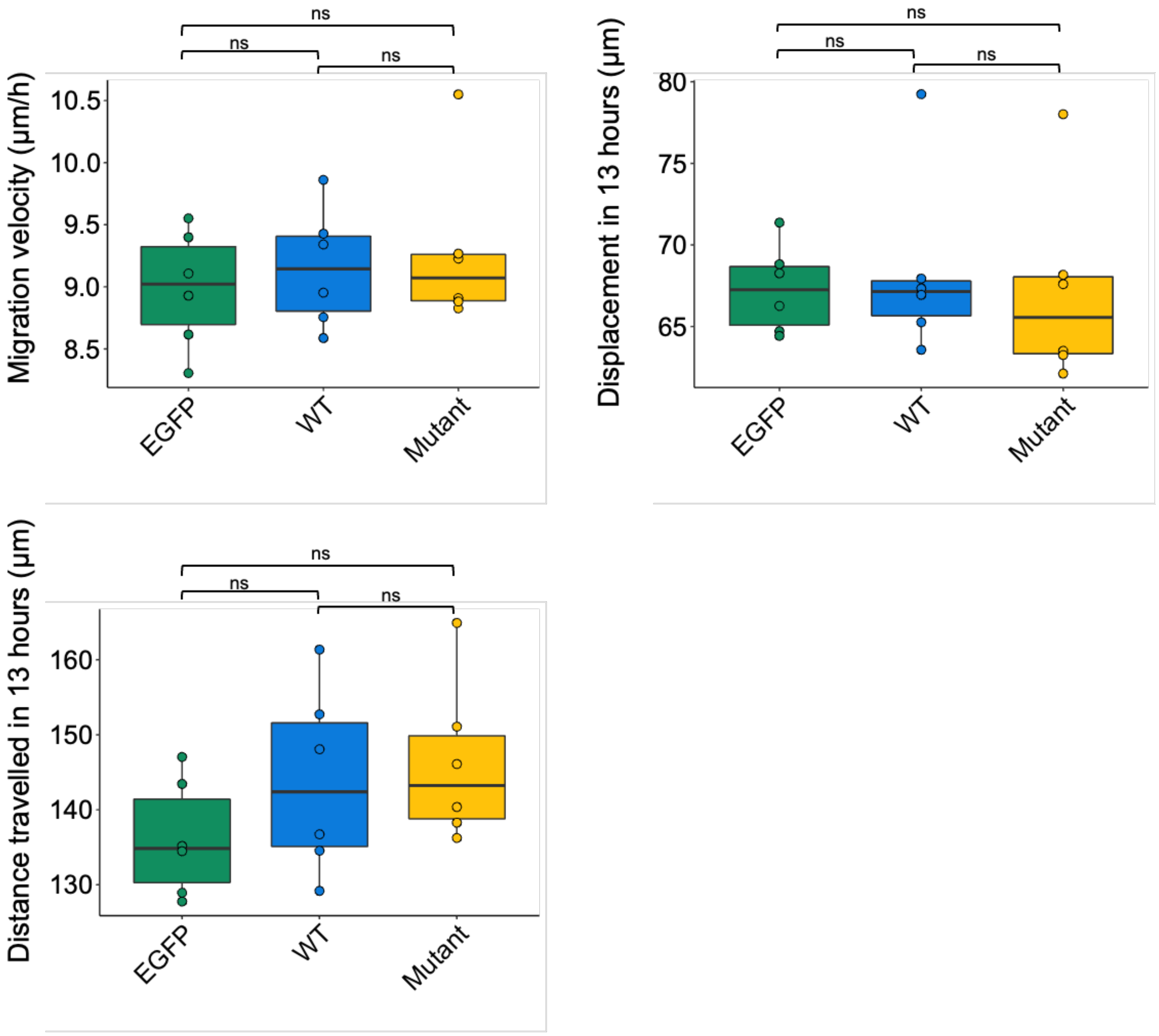

**Supplementary Figure 9. Flag-GEF18 expression in stable cell lines**

Expression of Flag-GEF18 in stable cell lines was quantified using Western blot and normalized to GAPDH expression. Significance was evaluated using a Kruskal-Wallis test followed by a post hoc Tukey test (****p<0.0001, ***p<0.001, **p<0.01, *p<0.05).

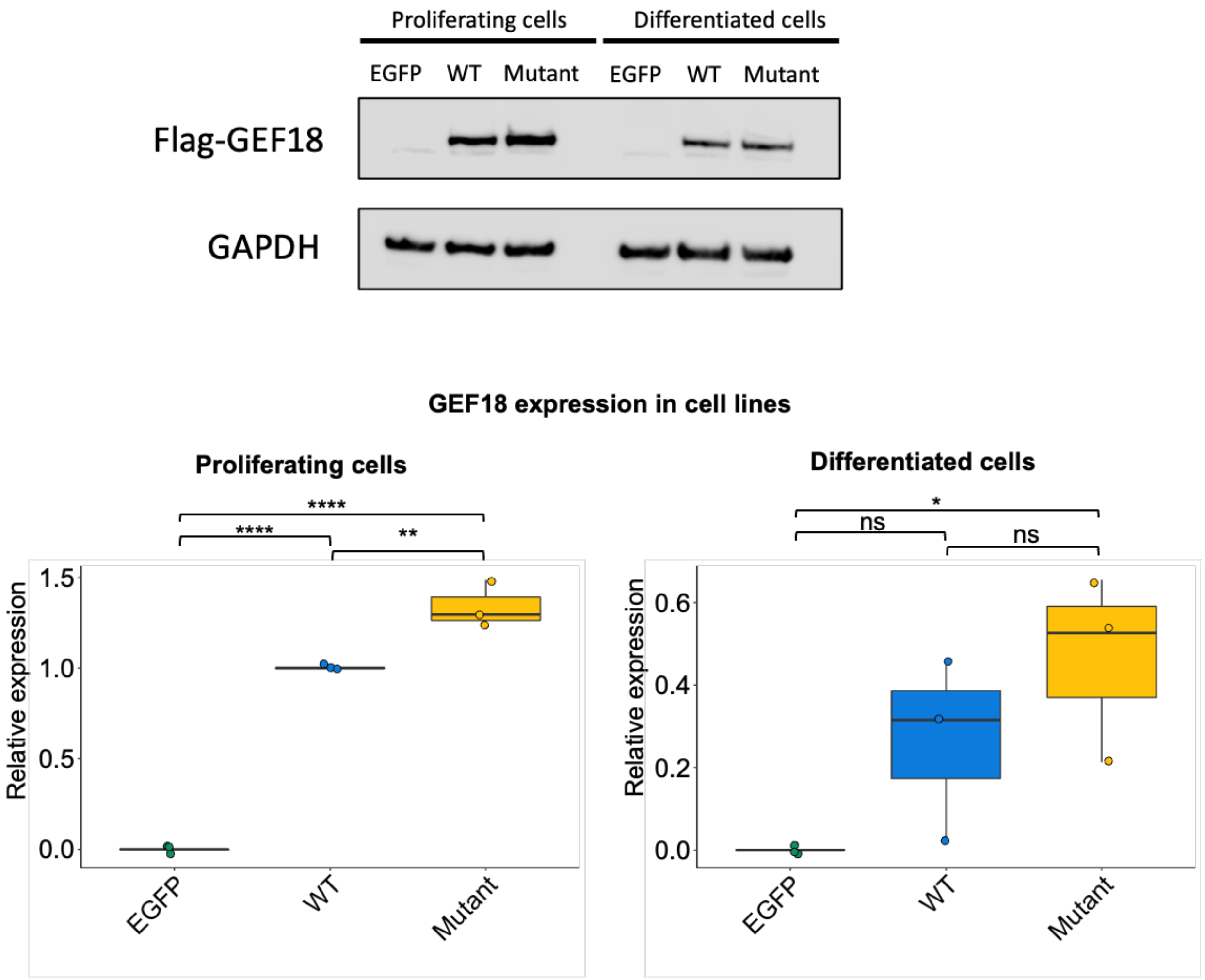

**Supplementary Tables**

**Supplementary Table 1. Laboratory assays significantly different between DKD clusters.** Labs used as input for clustering were compared between clusters using a t-test. The false discovery rate was computed using the Benjamini-Hochberg method.

| **Lab** | **Cluster M, Mean (SD)** | **Cluster S, Mean (SD)** | **p-value** | **FDR** |
|---|---|---|---|---|
| Blood Urea Nitrogen | 23.0 (7.7) | 27.0 (9.4) | 4.00E-13 | 2.00E-11 |
| Red Cell Distribution Width | 14.9 (1.4) | 15.4 (1.5) | 2.00E-08 | 3.00E-07 |
| Mean Corpuscular Hemoglobin Concentration | 33.4 (0.6) | 33.2 (0.67) | 1.00E-07 | 2.00E-06 |
| Blood Calcium | 9.4 (0.49) | 9.2 (0.52) | 1.00E-06 | 1.00E-05 |
| Blood Creatinine | 1.3 (0.37) | 1.4 (0.4) | 1.00E-06 | 1.00E-05 |
| Blood Albumin | 3.9 (0.41) | 3.8 (0.45) | 3.00E-05 | 2.00E-04 |
| Serum Sodium | 139.3 (2) | 138.8 (2) | 1.00E-04 | 7.00E-04 |
| Low Density Lipoprotein | 91.9 (24) | 85.8 (24) | 3.00E-04 | 0.001 |
| Mean Corpuscular Hemoglobin | 29.7 (1.9) | 29.4 (2) | 4.00E-04 | 0.002 |
| Total Protein in Blood | 7.1 (0.54) | 7.0 (0.59) | 6.00E-04 | 0.002 |
| Monocyte Percentage | 7.7 (1.7) | 8.0 (1.9) | 8.00E-04 | 0.003 |
| Blood Potassium | 4.3 (0.31) | 4.4 (0.31) | 0.001 | 0.004 |
| Hemoglobin | 11.8 (1.5) | 11.5 (1.5) | 0.002 | 0.005 |
| Lymphocyte Percentage | 23.6 (7.7) | 22.0 (7.9) | 0.002 | 0.005 |
| Serum Chloride | 102.5 (2.5) | 102.2 (2.8) | 0.005 | 0.01 |
| Mean Corpuscular Volume | 88.9 (4.9) | 88.4 (5.3) | 0.006 | 0.01 |
| Hematocrit | 35.8 (4.4) | 35.0 (4.7) | 0.02 | 0.04 |

**Supplementary Table 2. Differences in HPO terms between clinical clusters.** HPO terms with significant difference (FDR <0.05) in prevalence between clinical clusters are provided. HPO terms were extracted from clinical notes available in the EHR for each patient.

| HPO Term | Cluster M, n (%) | Cluster S, n (%) | p-value | FDR |
|---|---|---|---|---|
| Hypervolemia | 188 (19) | 127 (33) | 3.85E-07 | 0.000116283 |
| Renal insufficiency | 341 (35) | 194 (50) | 8.46E-07 | 0.000127769 |
| Abnormality of the tricuspid valve | 196 (20) | 126 (32) | 3.01E-06 | 0.000287119 |
| Tricuspid regurgitation | 194 (20) | 125 (32) | 3.80E-06 | 0.000287119 |
| Abnormality of the nasal cavity | 252 (26) | 58 (15) | 8.51E-06 | 0.000514075 |
| Obesity | 498 (51) | 149 (38) | 1.50E-05 | 0.000753483 |
| Abnormality of the nasal mucosa | 235 (24) | 54 (14) | 1.95E-05 | 0.000841178 |
| Rhinitis | 232 (24) | 54 (14) | 2.64E-05 | 0.000885658 |
| Pleural effusion | 341 (35) | 185 (47) | 2.78E-05 | 0.000885658 |
| Arterial calcification | 381 (39) | 201 (52) | 3.70E-05 | 0.000885658 |
| Peripheral arterial calcification | 381 (39) | 201 (52) | 3.70E-05 | 0.000885658 |
| Abnormality of the pulmonary vasculature | 278 (29) | 157 (40) | 3.81E-05 | 0.000885658 |
| Abnormality of pulmonary circulation | 278 (29) | 157 (40) | 3.81E-05 | 0.000885658 |
| Pulmonary opacity | 370 (38) | 195 (50) | 5.91E-05 | 0.001274106 |
| Hyperkalemia | 248 (26) | 142 (36) | 8.79E-05 | 0.00176874 |
| Atrial fibrillation | 197 (20) | 118 (30) | 0.000118071 | 0.002228588 |
| Chronic kidney disease | 490 (50) | 241 (62) | 0.000150249 | 0.002669135 |
| Coronary artery calcification | 200 (21) | 118 (30) | 0.000171535 | 0.00287798 |

| | | | | |
|---|---|---|---|---|
| Low back pain | 443 (46) | 135 (35) | 0.000214858 | 0.003415119 |
| Leukocytosis | 196 (20) | 115 (29) | 0.000267842 | 0.004044416 |
| Vascular calcification | 457 (47) | 226 (58) | 0.000316568 | 0.004529619 |
| Mitral regurgitation | 221 (23) | 126 (32) | 0.000338071 | 0.004529619 |
| Neck pain | 347 (36) | 100 (26) | 0.000344971 | 0.004529619 |
| Peripheral arterial stenosis | 581 (60) | 273 (70) | 0.000408561 | 0.005141059 |
| Arterial tortuosity | 223 (23) | 126 (32) | 0.00045247 | 0.005465836 |
| Abnormality of the aortic valve | 212 (22) | 121 (31) | 0.000475403 | 0.005521989 |
| Inflammation of the large intestine | 179 (18) | 106 (27) | 0.000513903 | 0.005748097 |
| Urinary incontinence | 257 (26) | 69 (18) | 0.000566998 | 0.006115476 |
| Supraventricular arrhythmia | 300 (31) | 158 (41) | 0.000767051 | 0.007987908 |
| Aortic tortuosity | 218 (22) | 121 (31) | 0.001110045 | 0.011174458 |
| Pustule | 385 (40) | 119 (31) | 0.001887652 | 0.018389386 |
| Primary atrial arrhythmia | 289 (30) | 150 (38) | 0.002078221 | 0.019613208 |
| Abnormality of the immune system | 336 (35) | 170 (44) | 0.002345125 | 0.021303652 |
| Congestive heart failure | 308 (32) | 158 (41) | 0.002398424 | 0.021303652 |
| Benign genitourinary tract neoplasm | 315 (32) | 94 (24) | 0.002606367 | 0.021864526 |
| Neoplasm of the genitourinary tract | 315 (32) | 94 (24) | 0.002606367 | 0.021864526 |
| Nonproductive cough | 340 (35) | 104 (27) | 0.003244732 | 0.025989907 |
| Headache | 528 (54) | 177 (45) | 0.003270253 | 0.025989907 |
| Abnormality of ion homeostasis | 392 (40) | 191 (49) | 0.003673966 | 0.028007946 |
| Microcytic anemia | 179 (18) | 100 (26) | 0.003709662 | 0.028007946 |
| Sleep disturbance | 489 (50) | 162 (42) | 0.003947794 | 0.029078872 |

| | | | | |
|---|---|---|---|---|
| Overweight | 617 (63) | 215 (55) | 0.004686902 | 0.033701058 |
| Arthralgia of the hip | 384 (40) | 122 (31) | 0.005210938 | 0.035324591 |
| Hip pain | 384 (40) | 122 (31) | 0.005210938 | 0.035324591 |
| Pneumothorax | 510 (52) | 237 (61) | 0.005608608 | 0.035324591 |
| Atelectasis | 504 (52) | 235 (60) | 0.005616044 | 0.035324591 |
| Abnormality of female internal genitalia | 408 (42) | 132 (34) | 0.00582424 | 0.035324591 |
| Abnormal morphology of the left ventricle | 293 (30) | 148 (38) | 0.005894341 | 0.035324591 |
| Gastrointestinal hemorrhage | 180 (19) | 99 (25) | 0.005911849 | 0.035324591 |
| Thoracic hypoplasia | 249 (26) | 129 (33) | 0.006068068 | 0.035324591 |
| Abnormality of cardiac atrium | 381 (39) | 185 (47) | 0.006170886 | 0.035324591 |
| Agitation | 214 (22) | 114 (29) | 0.006189205 | 0.035324591 |
| Pelvic mass | 379 (39) | 121 (31) | 0.006199349 | 0.035324591 |
| Insomnia | 353 (36) | 111 (28) | 0.006492061 | 0.036307452 |
| Abnormality of the atrioventricular valves | 332 (34) | 164 (42) | 0.007355948 | 0.040390841 |
| Arterial stenosis | 196 (20) | 105 (27) | 0.007509086 | 0.040495429 |
| Cardiomegaly | 435 (45) | 206 (53) | 0.008199581 | 0.042640176 |
| Pruritus | 494 (51) | 167 (43) | 0.008303805 | 0.042640176 |
| Knee pain | 491 (51) | 166 (43) | 0.008330366 | 0.042640176 |
| Hypotension | 351 (36) | 171 (44) | 0.009568355 | 0.048160718 |

**Supplementary Table 3. Distribution of mean vitals across DKD clusters.** Mean vitals used as input for clustering were compared between clusters using a t-test. The false discovery rate was computed using the Benjamini-Hochberg method.

| **Measurement** | **Cluster M, Mean (SD)** | **Cluster S, Mean (SD)** | **P Value** | **FDR** |
|---|---|---|---|---|
| Respiratory Rate (breaths per min) | 17.86 (1.5) | 18.41 (1.8) | 1.00E-07 | 8.00E-07 |
| Weight (lbs) | 186.92 (47) | 175.06 (41) | 5.00E-06 | 2.00E-05 |
| Pulse Oxygen | 97.3 (1.2) | 97.12 (1.6) | 0.05 | 0.1 |
| Diastolic Blood Pressure (mmHg) | 70.92 (6.7) | 71.07 (7.2) | 0.7 | 0.8 |
| Height | 64.23 (4.1) | 64.14 (4.4) | 0.7 | 0.8 |
| Heart Rate (beats per min) | 76.75 (9.3) | 77.17 (9.8) | 0.5 | 0.8 |
| Systolic Blood Pressure (mmHg) | 135.97 (14) | 135.69 (15) | 0.8 | 0.8 |

**Supplementary Table 4. GWAS summary statistics for cluster M**. SNPs significantly associated with cluster M are provided (P < 5x10E-6). A logistic regression model was fit adjusted for age, sex, and the top 10 genetic principal components.

| | | | | | | | Cases (N) | | | Controls (N) | | |
|---|---|---|---|---|---|---|---|---|---|---|---|---|
| Chr | ID | OR | P | Gene | Protein change | SNP Type | REF/REF | REF/ALT | ALT/ALT | REF/REF | REF/ALT | ALT/ALT |
| chr1 | rs149369913 | 12.093 | 6.90E-07 | PADI2 | R619C | Missense | 939 | 5 | 0 | 27212 | 65 | 0 |
| chr19 | rs58142912 | 10.18 | 2.07E-07 | CBARP | A229T | Missense | 934 | 10 | 0 | 27195 | 80 | 2 |

**Supplementary Table 5. GWAS summary statistics for cluster S**. SNPs significantly associated with cluster S are provided (P < 5x10E-6). A logistic regression model was fit adjusted for age, sex, and the top 10 genetic principal components.

| | | | | | | | Cases (N) | | | Controls (N) | | |
|---|---|---|---|---|---|---|---|---|---|---|---|---|
| Chr | ID | OR | P | Gene | Protein change | SNP Type | REF/REF | REF/ALT | ALT/ALT | REF/REF | REF/ALT | ALT/ALT |
| chr19 | rs117824875 | 7.651 | 9.56E-08 | ARHGEF18 | A1033T | Missense | 367 | 8 | 0 | 27086 | 191 | 0 |
| chr22 | rs143008696 | 12.8 | 1.03E-07 | IL17RA | T51M | Missense | 370 | 5 | 0 | 27194 | 80 | 0 |
| chr2 | rs115032706 | 9.652 | 2.13E-07 | ASIC4 | A37V | Missense | 369 | 6 | 0 | 27159 | 118 | 0 |
| chr14 | rs370465761 | 10.51 | 6.51E-07 | GALNT16 | A549A | Synonymous | 367 | 8 | 0 | 27231 | 46 | 0 |
| chr12 | rs149467228 | 4.928 | 9.81E-07 | KRT5 | A357A | Synonymous | 365 | 9 | 1 | 27073 | 203 | 1 |
| chr12 | rs148757217 | 6.211 | 1.04E-06 | KRT83 | C423S | Missense | 366 | 9 | 0 | 27131 | 143 | 0 |
| chr5 | rs151112453 | 8.521 | 1.21E-06 | CMYA5 | S1725S | Synonymous | 369 | 0 | 0 | 27189 | 0 | 0 |
| chr3 | rs139311375 | 8.968 | 1.26E-06 | NME9 | N194K | Missense | 369 | 6 | 0 | 27204 | 73 | 0 |
| chr14 | rs199645167 | 9.781 | 1.28E-06 | ZNF839 | D895N | Missense | 369 | 6 | 0 | 27220 | 57 | 0 |
| chr1 | rs143442822 | 11698 | 1.34E-06 | IGSF9 | P51P | Synonymous | 373 | 2 | 0 | 27171 | 69 | 2 |
| chr12 | rs141404620 | 5.702 | 1.46E-06 | DCP1B | S350S | Synonymous | 365 | 10 | 0 | 27160 | 117 | 0 |

| | | | | | | | | | | | | |
|---|---|---|---|---|---|---|---|---|---|---|---|---|
| chr11 | rs76286372 | 11002 | 1.56E-06 | KCNK7 | A120A | Synonymous | 371 | 0 | 0 | 26952 | 0 | 0 |
| chr22 | rs140221307 | 9.815 | 1.60E-06 | IL17RA | W320R | Missense | 370 | 5 | 0 | 27177 | 99 | 0 |
| chr15 | rs149423703 | 7.673 | 2.04E-06 | ATP10A | A1250A | Synonymous | 366 | 7 | 0 | 27163 | 78 | 0 |
| chr6 | rs142498910 | 12.31 | 2.23E-06 | MDFI | P98R | Missense | 366 | 0 | 0 | 27083 | 0 | 0 |
| chr19 | rs113812503 | 3.373 | 2.69E-06 | ZIM2 | T339M | Missense | 358 | 17 | 0 | 26755 | 517 | 5 |
| chr8 | rs117256395 | 6.643 | 2.71E-06 | COL22A1 | P1086A | Missense | 368 | 7 | 0 | 27132 | 130 | 0 |
| chr2 | rs201144938 | 8467 | 2.97E-06 | SP140 | E797E | Synonymous | 331 | 44 | 0 | 23704 | 3549 | 13 |
| chr18 | rs140794493 | 3.74 | 3.37E-06 | CNDP2 | V352V | Synonymous | 361 | 0 | 0 | 27066 | 0 | 0 |
| chr2 | rs115833267 | 9.121 | 3.81E-06 | EPB41L5 | Q654L | Missense | 370 | 5 | 0 | 27179 | 98 | 0 |
| chr13 | rs143923184 | 7539 | 3.83E-06 | ENOX1 | T206T | Synonymous | 373 | 2 | 0 | 27186 | 88 | 1 |
| chr1 | rs12093154 | 1.624 | 4.11E-06 | C1QTNF12 | A180V | Missense | 274 | 89 | 12 | 22183 | 4780 | 307 |
| chr1 | rs76074441 | 5.627 | 4.34E-06 | GPA33 | V19I | Missense | 366 | 9 | 0 | 27179 | 98 | 0 |
| chr7 | rs72657364 | 7505 | 4.39E-06 | DNAH11 | I2682F | Missense | 373 | 2 | 0 | 27060 | 214 | 1 |
| chr7 | rs76996735 | 7646 | 4.59E-06 | ASNS | A6A | Synonymous | 364 | 11 | 0 | 26686 | 582 | 5 |
| chr3 | rs146581075 | 9.977 | 4.83E-06 | A4GNT | Y312C | Missense | 370 | 5 | 0 | 27220 | 57 | 0 |

**Supplementary Table 6. Association of rs117824875 in UK Biobank diabetic patients**. DKD was defined as individuals with a history of T2D and eGFR <60 at enrollment. P-value was estimated using a logistic regression model adjusted for age, sex, and five genetic principal components.

| Outcome | OR (95% CI) | p-value | N cases (carriers) | N controls (carriers) |
|---|---|---|---|---|
| DKD (T2D + eGFR <60) | 2.4 (1.02 - 5.42) | 0.04 | 680 (13) | 10090 (121) |

**Supplementary Videos**

Supplementary video 1: Representative live cell TIRF time-lapse video of $GEF18^{WT}$ mClover-fusion cell over a period of 6 hours under proliferative conditions

Supplementary video 2: Mean fluorescence intensity based polar plot showing changes in GEF18 subcellular localization in proliferating $GEF18^{WT}$ mClover-fusion cells

Supplementary video 3: Representative live cell TIRF time-lapse video of $GEF18^{MT}$ mClover-fusion cell over a period of 6 hours under proliferative conditions

Supplementary video 4: Mean fluorescence intensity based polar plot showing changes in GEF18 subcellular localization in proliferating $GEF18^{MT}$ mClover-fusion cells

Supplementary video 5: Representative live cell TIRF time-lapse video of $GEF18^{WT}$ mClover-fusion cell over a period of 6 hours under differentiated conditions

Supplementary video 6: Mean fluorescence intensity based polar plot showing changes in GEF18 subcellular localization in terminally differentiated $GEF18^{WT}$ mClover-fusion cells

Supplementary video 7: Representative live cell TIRF time-lapse video of $GEF18^{MT}$ mClover-fusion cell over a period of 6 hours under differentiated conditions

Supplementary video 8: Mean fluorescence intensity based polar plot showing changes in GEF18 subcellular localization in terminally differentiated $GEF18^{MT}$ mClover-fusion cells
