## Supplementary Note 1 for "Deep learning on electronic medical records identifies distinct subphenotypes of diabetic kidney disease driven by genetic variations in the *Rho* pathway"

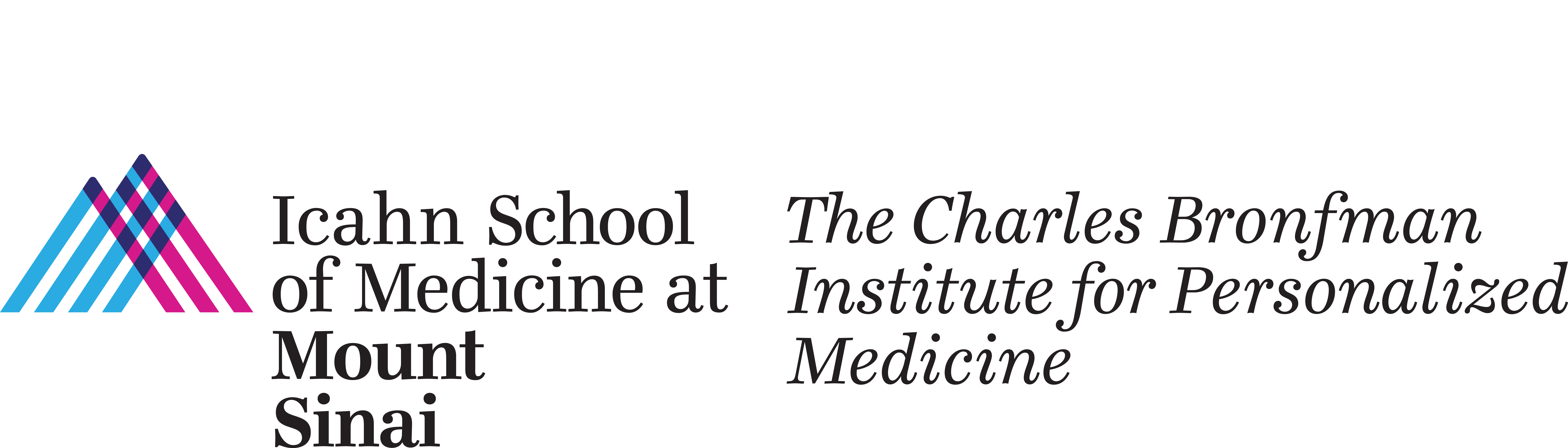

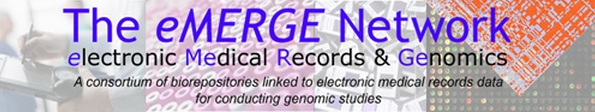

**Phenotyping Algorithm for**

**Chronic Kidney Disease (CKD)**

**Associated with Type 2 Diabetes Mellitus**

**Or Hypertension**

v1.7 March 2014

**CKD Definitions, Staging and Disease Burden**

| **Definition** | |
| --- | --- |
| Functional criteria | eGFR <60 mL/min per 1·73 m^2^ for >3 months |
| Structural criteria | Kidney damage for >3 months (albuminuria is the most common marker of kidney damage and is also associated with rapid progression) |
| **Staging** | |
|  | eGFR categories (mL/min per 1·73 m^2^) and related terms: G1 ≥90 (normal or high); G2 60–89 (mildly decreased); G3a 45–59 (mildly to moderately decreased); G3b 30–44 (moderately to severely decreased); G4 15–29 (severely decreased); G5 <15 (kidney failure) |
| **Burden** | |
| Prevalence | ∼10% of adults (from 4% at 20–39 years to 47% at ≥70 years in the USA) |
| Annual incidence | ∼1% in middle age; twice as frequent in black compared with white populations |
| Lifetime cumulative incidence | ∼50% for chronic kidney disease and ∼2% in white and ∼7% in black populations for end-stage renal disease |
|  | *eGFR=estimated glomerular filtration rate.* |
|  | *In the absence of evidence of kidney damage, eGFR category G1 or G2 do not fulfill the criteria for chronic kidney disease.* |

The two main causes of chronic kidney disease are **diabetes and high blood pressure, which are responsible for up to two-thirds of the cases**.

**Diabetes/Hypertension-Associated CKD (eGFR < 60 ml/min/1.73m^2^)**

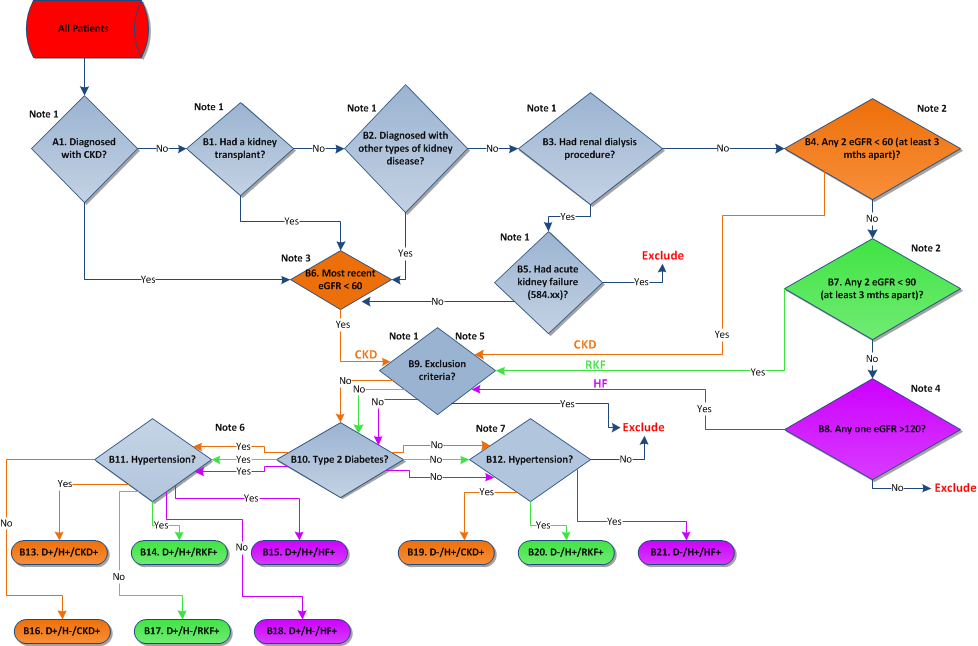

For detailed information on each box, please see the respective Note

**NOTE 1:**

**ICD-9 Diagnosis and Procedure codes applied in algorithm**

| **A1** |  |
| --- | --- |
| 585.xx | Chronic kidney disease |
| **B1** |  |
| 55.6x | Transplant of kidney (procedure) |
| V42.0 | Organ or tissue replaced by kidney transplant |
| **B2** |  |
| 250.4x | Diabetes with renal manifestations |
| 403.xx | Hypertensive chronic kidney disease |
| 404.xx | Hypertensive heart and chronic kidney disease |
| **B3** |  |
| V45.1 | Renal dialysis status |
| V56.xx | Encounter for dialysis and dialysis catheter care |
| 996.73 | Complications of renal dialysis |
| 38.95 | Venous catheter for renal dialysis |
| **B5** |  |
| 584.xx | Acute kidney failure |
| **B9** |  |
| 042.xx-044.xx | Human immunodeficiency virus (HIV) infection |
| 282.6 | Sickle cell disease |
| 581.xx | Nephrotic syndrome |
| 582.xx | Chronic glomerulonephritis |
| 583.xx | Nephritic and nephropathy |
| 446.xx | Polyarteritis nodosa and allied conditions |
| 447.6 | Vasculitis |
| 753.xx | Renal agenesis and dysgenesis |

ICD 10 :

A1 : N18

B1 : Z94.0,0TS00ZZ,0TS10ZZ,0TY00Z0,0TY00Z1,0TY00Z2,0TY10Z0,0TY10Z1,0TY10Z2

B2 : E11.29,E10.29,E11.21,E11.65,E10.21,E10.65,I12.9,I12.0,I13

B3 : T82.818A,T82.828A,T82.838A,T82.848A,T82.858A,T82.868A,T82.898A,Z99.2,Z91.15,Z49, 05HY33Z,06HY33Z

B5 : N17

B9 : B20,D57,N02.2,N08,N04,N03,N05,N17.1,N17.2,M30.0,M30.3,M31.0,M31.2,M31.30,M31.6,

M31.1,M31.4,I77.6,Q60.2,Q60.5,Q61.00,Q61.9,Q61.01,Q61.3,Q61.2,Q61.19,Q61.4,Q61.5,Q61.02,Q61.8,Q62.39,

Q62.11,Q62.12,Q62.31,Q62.10,Q63.0,Q63.1,Q63.2,Q63.3,Q63.8,Q62.4,Q62.5,Q62.61,Q62.62,Q62.63,Q62.8,

Q64.10,Q64.19,Q64.2,Q64.31,Q64.32,Q64.33,Q64.39,Q64.4,Q64.5,Q64.6,Q64.71,Q64.73,Q64.74,Q64.75,Q64.79,Q64.9

**NOTES 2, 3, 4 eGFR criteria for CKD definition:**

**Calculation of estimated glomerular filtration rate (eGFR) and its application**

Various formats/formulas are used to estimated glomerular filtration rate (eGFR) based on measured serum creatinine lab results, age, gender, and race, leading to non-uniformity of eGFR results reported by clinical laboratories to the EHR. In this CDA, we overcome the apparent non-uniformity of eGFR lab results calculation and reporting in EHR by recalculating eGFR de novo with the CKD-EPI formula.

Ref: Levey AS, Stevens LA, et al. A New Equation to Estimate Glomerular Filtration Rate. [Ann Intern Med. 2009; 150:604-612.](http://www.ncbi.nlm.nih.gov/sites/entrez?db=pubmed&term=19414839)

The CKD-EPI creatinine equation is based on the same four variables as the MDRD Study equation, but uses a 2-slope “spline” to model the relationship between estimated GFR and serum creatinine, and a different relationship for age, sex and race. The equation was reported to perform better and with less bias than the MDRD Study equation, especially in patients with higher GFR. This results in reduced misclassification of CKD.

**CKD-EPI equation**

| **Race** | **Sex** | **Serum Creatinine (SCr) threshold (mg/dl)** | **Equation** |
| --- | --- | --- | --- |
| Black | Female | <=0.7 | GFR = 166 × (SCr/0.7) ^-0.329^ × (0.993)^Age^ |
| Black | Female | >0.7 | GFR = 166 × (SCr/0.7) ^-1.209^ × (0.993)^Age^ |
| Black | Male | <=0.9 | GFR = 163 × (SCr/0.9) ^-0.411^ × (0.993)^Age^ |
| Black | Male | >0.9 | GFR = 163 × (SCr/0.9) ^-1.209^ × (0.993)^Age^ |
| White or other | Female | <=0.7 | GFR = 144 × (SCr/0.7) ^-0.329^ × (0.993)^Age^ |
| White or other | Female | >0.7 | GFR = 144 × (SCr/0.7) ^-1.209^ × (0.993)^Age^ |
| White or other | Male | <=0.9 | GFR = 141 × (SCr/0.9) ^-0.411^ × (0.993)^Age^ |
| White or other | Male | >0.9 | GFR = 141 × (SCr/0.9) ^-1.209^ × (0.993)^Age^ |

**Exclusion criteria:** We did not allow patients that had serum creatinine tests on consecutive days or within the same day as we assumed these were ‘inpatients’. For patients that had duplicate serum creatinine tests meaning they had two tests at the same date/time but different values we kept the test with the maximum value in the algorithm.

**NOTE 2:**

For B4 and B7 case patients, we used any 2 eGFR results among all their records which had at least a 90 day window between them and evaluated them under the thresholds below.  Note that patients with only one eGFR<60 and at least one 60<=eGFR<90 three months apart are classified as RKF cases.   For B4 control patients we simply required that they **not** have an eGFR <90 ml/min/1.73m^2^ **ever**; and for B8 control patients that they **not** have an eGFR >120 ml/min/1.73m^2^ **ever**.  For controls, if there are no eGFR results available, this was considered “No” for B4 and B8; those patients are eligible (input into B9) to potentially be included as controls.

- Chronic Kidney Disease CKD threshold < 60 ml/min/1.73m^2^ (box B4 cases)
- Reduced Kidney Function (RKF) threshold < 90 ml/min/1.73m^2^ (box B7 cases and B4 controls–identification of RKF patients is required for exclusion from CKD controls).
- Hyperfiltration (HF) threshold > 120 ml/min/1.73m^2^ (box B8 cases and B8 controls – identification of HF patients is required for exclusion from CKD controls).

**NOTE 3:**

Patient count for output of box B6 was determined by including patients with a recorded diagnosis code for CKD (see A1) that had their last eGFR result lower than 60.

**NOTE 4:**

Patient count for output of box B8 was determined by including patients that had at least one eGFR result value higher than 120 to identify patients with increased kidney function (hyperfiltration). Because hyperfiltration can indicate an often unrecognized early stage of kidney disease, including diabetic and hypertensive CKD, identification of patients is required for exclusion from CKD Controls (see CKD Controls algorithm, box B8).

**NOTE 5:**

The ICD-9 codes in Note 1 and the following terms found in observation reports by the physician were used as exclusion criteria for B9 (see end of document for Code detail):

- [HIVAN/HIV associated nephropathy]
- Congenital (within 2 words of) kidney(s)
- [APKD/Adult polycystic kidney disease]
- Sickle Cell Disease
- IgA Nephropathy
- Nephrotic Syndrome
- Nephritic Syndrome
- Glomerulonephritis
- Glomerulosclerosis
- Lupus Nephritis
- Wegener's granulomatosis
- Goodpasture's syndrome

**NOTE 6:**

For box B10, we ran the output of ‘B9->No’ patients against the Modified Northwestern Type 2 Diabetes CDA (Page 9). The yes-cases from box B10 are run against the IPM Hypertension CDA (Page 11) in box B11, and the no-cases from box B10 are run against box B12:

Yes-cases from box B11 are diabetic and hypertensive (DH) cases and are grouped as follows:

- B13 = DH-CKD - Cases of CKD Stage III or higher if yes-cases from box B4 or B6;
- B14 = DH-RKF - Patients with reduced kidney function: if yes-cases from box B7;
- B15 = DH-HF - Patients with glomerular hyperfiltration: if yes-cases from box B8.

No-cases from box B11 are non-hypertensive diabetic (D) cases and are grouped as follows:

- B16 = D-CKD - Cases of CKD Stage III or higher if yes-cases from box B4 or B6;
- B17 = D-RKF - Patients with reduced kidney function: if yes-cases from box B7;
- B18 = D-HF - Patients with glomerular hyperfiltration: if yes-cases from box B8.

**NOTE 7:**

The no-cases from box B12 are excluded. The yes-cases from box B12 are non-diabetic hypertensive (H) cases and are grouped as follows:

- B19 = H-CKD - Cases of CKD Stage III or higher if yes-cases from box B4 or B6;
- B20 = H-RKF - Patients with reduced kidney function: if yes-cases from box B7;
- B21 = H- HF - Patients with glomerular hyperfiltration: if yes-cases from box B8.

**CKD CDA TEXT SEARCH:**

**Descriptors for exclusion criterion (Box B9):**

- HIVAN/HIV associated nephropathy]
- Congenital (within 2 words of) kidney(s)
- [APKD/Adult polycystic kidney disease]
- Sickle Cell Disease
- IgA Nephropathy
- Nephrotic Syndrome
- Nephritic Syndrome
- Glomerulonephritis
- Glomerulosclerosis
- Lupus Nephritis
- Wegener's granulomatosis
- Goodpasture's syndrome

**The code picks up all the patients who have the above mentioned terms in the progress notes as long as they are not preceded or followed by “negative/no/not/absence” (within 3 words for nephropathy, 2 words for congenital, apkd & sickle) or family history related terms (50 characters prior to 50 characters after) which are detailed below.**

[DESCRIPTORS]

-Without ‘**negative/no/not/ absence**’ preceded within n words (depending on the notes)

-Without ‘**sister/brother/mother/father/grandfather/grandmother/family/ grandson/granddaughter/son/daughter**’ within 50 characters before or 50 characters after keyword

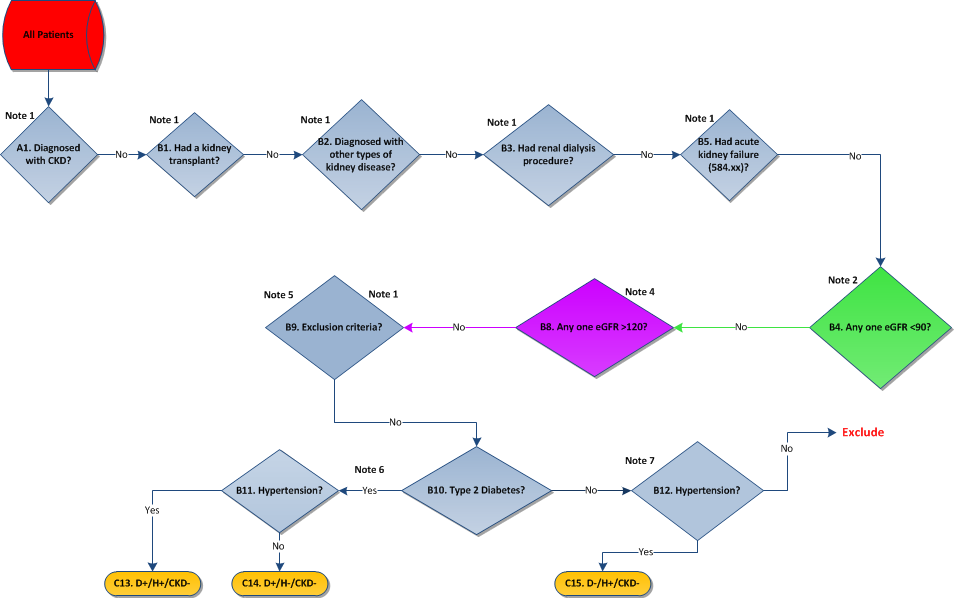

**Diabetes/Hypertension-Associated CKD Controls**

**Controls**

CKD Control groups:

C13 = yes-records from box B11 are diabetic and hypertensive (DH-Control) controls for DH-CKD cases (box B13).

C14 = no-records from box B11 are diabetic (D-Controls) controls without hypertension for D-CKD cases (box B16).

C15 = yes-records from box B12 are hypertensive (H-Controls) controls without diabetes for H-CKD cases (box B19).

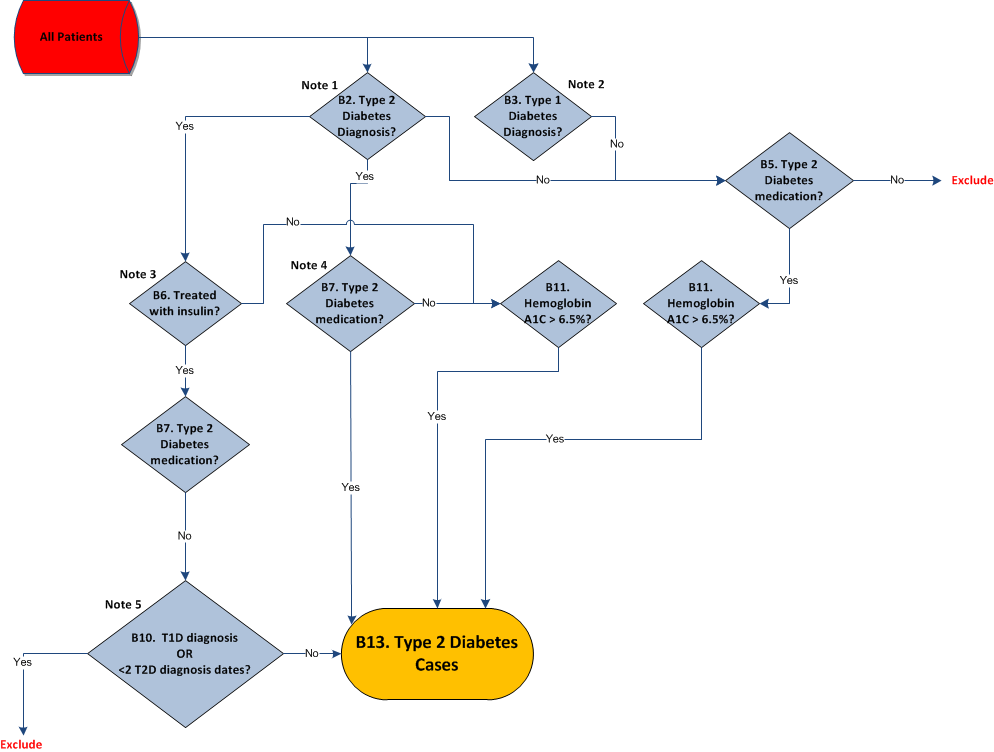
**NOTE 1. All T2 Diabetes ICD-9 Codes:**

**Type 2 Diabetes Cases**

(modified from Northwestern eMERGE Phase 1 CDA)

-250.00, 250.10, 250.20, 250.30, 250.40, 250.50, 250.60, 250.70, 250.80, 250.90, 250.02, 250.12, 250.22, 250.32, 250.42, 250.52, 250.62, 250.72, 250.82, 250.92

**NOTE 2. All T1 Diabetes ICD-9 Codes:**

-250.01, 250.11, 250.21, 250.31, 250.41, 250.51, 250.61, 250.71, 250.81, 250.91, 250.03, 250.13, 250.23, 250.33, 250.43, 250.53, 250.63, 250.73, 250.83, 250.93

**NOTE 3. Including all drugs with active ingredient insulin, list of terms used to search for associated patients:**

-“APIDRA”, “APIDRA SOLOSTAR”, “EXUBERA”, “HUMALOG”, “HUMALOG KWIKPEN”, “HUMALOG MIX 50/50”, “HUMALOG MIX 50/50 KWIKPEN”, “HUMALOG MIX 50/50 PEN”, “HUMALOG MIX 75/25”, “HUMALOG MIX 75/25 KWIKPEN”, “HUMALOG MIX 75/25 PEN”, “HUMALOG PEN”, “HUMULIN 50/50”, “HUMULIN 70/30”, “HUMULIN 70/30 PEN”, “HUMULIN BR”, “HUMULIN L”, “HUMULIN N”, “HUMULIN R”, “HUMULIN R PEN”, “HUMULIN U”, “ILETIN I”, “ILETIN II”, “INSULATARD NPH HUMAN”, “INSULIN”, “INSULIN INSULATARD NPH NORDISK”, “INSULIN NORDISK MIXTARD (PORK)”, “LANTUS”, “LENTARD”, “LENTE”, “LENTE ILETIN II”, “LENTE ILETIN II (PORK)”, “LENTE INSULIN”, “LEVEMIR”, “LEVEMIR”, “MIXTARD HUMAN 70/30”, “NOVOLIN 70/30”, “NOVOLIN L”, “NOVOLIN N”, “NOVOLIN R”, “NOVOLOG”, “NOVOLOG MIX 50/50”, “NOVOLOG MIX 70/30”, “NPH ILETIN I (BEEF-PORK)”, “NPH ILETIN II”, “NPH ILETIN II (PORK)”, “NPH INSULIN”, “NPH PURIFIED PORK ISOPHANE INSULIN”, “PROTAMINE ZINC & ILETIN I (BEEF-PORK)”, “PROTAMINE ZINC AND ILETIN II”, “PROTAMINE ZINC AND ILETIN II (PORK)”, “PROTAMINE ZINC INSULIN”, “REGULAR ILETIN II”, “REGULAR ILETIN II (PORK)”, “REGULAR INSULIN”, “REGULAR PURIFIED PORK INSULIN”, “SEMILENTE”, “SEMILENTE INSULIN”, “ULTRALENTE”, “ULTRALENTE INSULIN”, “VELOSULIN”, “VELOSULIN BR”, “VELOSULIN BR HUMAN”

**NOTE 4. List of terms used to search for all Type 2 Diabetes medications:**

"ACARBOSE", "ACTOPLUS MET", "ACTOPLUS MET XR", "ACTOS", "AMARYL", "AVANDAMET", "AVANDARYL", "AVANDIA", "DIABETA", "DUETACT", "FORTAMET", "GLIMEPIRIDE", "GLIPIZIDE", "GLIPIZIDE AND METFORMIN HYDROCHLORIDE", "GLIPIZIDE AND METFORMIN HYDROCHLORIDE", "GLUCOPHAGE", "GLUCOPHAGE XR", "GLUCOTROL", "GLUCOTROL XL", "GLUCOVANCE", "GLUMETZA", "GLYBURIDE", "GLYBURIDE (MICRONIZED)", "GLYBURIDE AND METFORMIN HYDROCHLORIDE", "GLYNASE", "JANUMET", "METAGLIP", "METFORMIN HYDROCHLORIDE", "MICRONASE", "PIOGLITAZONE", "PRANDIMET", "PRANDIN", "PRECOSE", "REPAGLINIDE", "RIOMET", "ROSIGLITAZONE", "ROSIGLITAZONE MALEATE", "ROSIGLITAZONE MALEATE; GLIMEPIRIDE", "ROSIGLITAZONE MALEATE; METFORMIN HYDROCHLORIDE"

**NOTE 5:**

To be excluded from having ‘< 2 T2 diagnosis dates’, patients must have 2 DISTINCT dates or more, that is not 2 diagnoses on the same day and not including null date values.

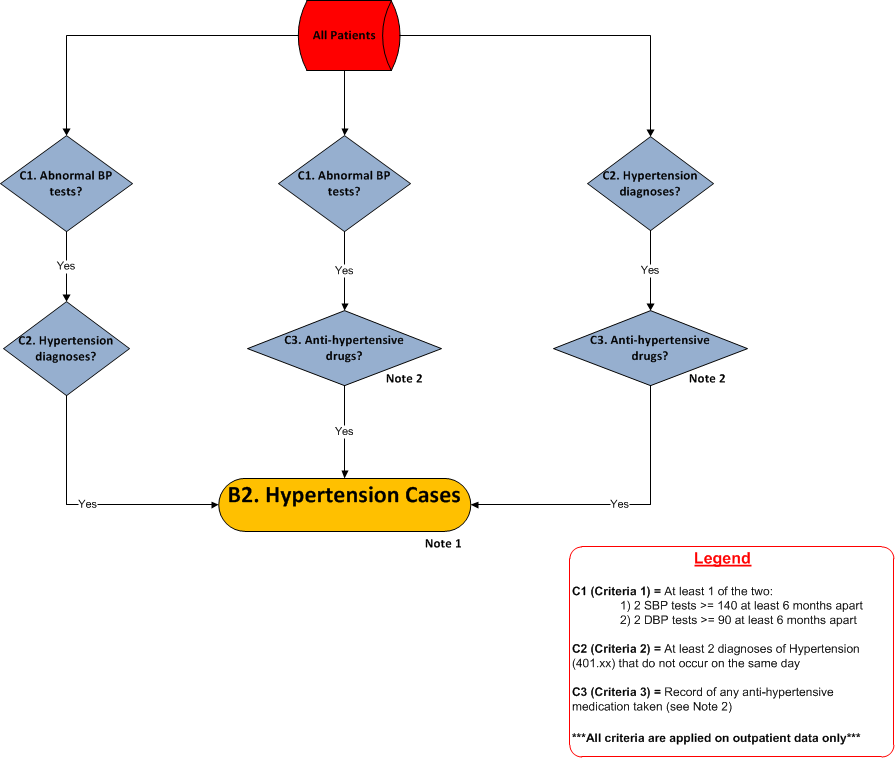
**NOTE 1:**

**Hypertension Cases**

The Hypertension cases total (B2) is assembled from the three major hypertension criteria: 2 Abnormal SBP or 2 abnormal DBP lab tests 6 months apart (C1), at least 2 Hypertension diagnoses with ICD-9 code 401.xx that are not on the same day (C2) and a record of taking any anti-hypertensive drug listed in Note 2 (C3). Essentially the B2 total box is made up of patients that have **any** 2 of the 3 major hypertension criteria by taking the union of unique patients from each criteria combination pair.

**NOTE 2:**

**Anti-Hypertensive Drugs**

**(Taken from eMERGE ‘Resistant Hypertension’ Network Phenotyping Algorithm)**

**Medication Classes:**

**Hydralazine**: Hydralazine (Apresazide  , bidil, apressoline)

***Minoxidil (****Loniten)*

***Renin antagonist****: aliskiren (Tekturna)*

**Central alpha agonists**: clonidine (catapres) (catapress); guanabenz; methyldopa; methyldopate

**ACEI/ARB**: candesartan (Atacand); Irbesartan (avapro); lisinopril (prinivil, zestril); trandolapril (Mavik, gopten, odrik); Losartan (cozaar); enalapril (enalaprilat); valsartan (diovan); telmisartan (Micardis); moexipril; quinapril (accupril); ramipril (altace); fosinopril (monopril); eprosartan; olmesartan (benicar); perindopril (Aceon); captopril (Capoten); benazepril (lotensin);

**Aldosterone antagonists**: spironolactone (Aldactone); eplerenone (inspra)

**Diuretics (count each instance as part of the same class, even if on more than one concurrently): Thiazide**: hydrochlorothiazide (Esidrix); indapamide (lozol, natrilix); cyclothiazide; chlorothiazide; chlorthalidone; bendroflumethiazide; benzothiazide; **K-sparing diuretics:** amiloride (midamor); triamterene (Dyrenium); ***Loop diuretics:*** *furosemide (lasix), torsemide (demadex), ethacrynic acid (ethacrynate, edecrin); bumetanide (bumex)*

**Note: the Diuretic combination meds now count as a single class:** Dyazide, Moduretic, Maxzide

**Alpha antagonists**: prazosin (minipress); doxazosin (cardura)

**Non-dihydro. CCBs**: verapamil (calan, covera, isoptin, verelan); diltiazem (dilt, tiazac, cardizem)

**Dihydro CCBs**: isradipine (Dynacirc); nicardipine; nifedipine (procardia); nisoldipine; felodipine (plendil); Amlodipine (norvasc, caduet); bepridil (vascor)

**Beta Blockers**: propranolol (inderal); metoprolol (toprol); labetalol (trandate); nadolol (corgard); esmolol (brevibloc); pindolol; penbutolol (levatol); Labetalol (Normodyne); atenolol (tenormin); carvedilol (coreg); bisoprolol (Zebeta);

***Thiazide/BB**: corzide , Tenoretic, lopressor HCT

***Thiazide/ACEI_ARB**: zestoretic, Avalide, hyzaar, uniretic, benicar HCT, accuretic, Teveten HCT, lotensin HCT; micardis HCT; atacand HCT; Diovan HCT; Monopril HCT

***Thiazide/aldosterone antagonist**: aldactazide
****Thiazide/Renin antagonist****: Tekturna HCT*

**Diabetes/Hypertension-Associated Chronic Kidney Disease (CKD)**

**Phenotyping Algorithm Validation Protocol**

The aim of this protocol is to provide performance statistics (sensitivity, specificity, PPV, NPV) for the eMERGE Phase II DH-CKD phenotyping algorithm.

1. Run IPM DH-CKD algorithm on clinical database, limiting to those that have data entered after January 1 2003.
2. Select 25 patients each at random from outputs D1, DH1 and H1 from the CKD case definition algorithm and outputs C1, C2 and C3 from the control definition algorithm. Total number of charts for review will be 150 (75 cases and 75 controls).
3. Patient list to be randomized and blinded (remove column B in recommended format below) prior to review
4. Physician reviewer to assign case/control status. Case status = 1, Control status = 0, Unknown/Other = 3 (please add text to describe anomaly). Please use the following format (removing column B, ‘Algorithm Status’, prior to physician review):

| **Patient ID** | **Algorithm Status** | **CKD** | **HTN** | **T2D** | **Text** |
| --- | --- | --- | --- | --- | --- |
| XXXXXX | D1 | 1 | 0 | 1 |  |
| XXXXXX | DH1 | 1 | 1 | 1 |  |
| XXXXXX | H1 | 1 | 1 | 0 |  |

5. Status should be based on the following criteria:
A. **CKD:**

- **Inclusion criteria** (if any present mark as 1, if none present mark as 0 (unless exclusion criteria apply – see below)):
- Documented diagnosis of CKD
- Renal transplant/Renal replacement therapy
- Evidence of consistent eGFR’s <60
- **Exclusion criteria** (if any present, mark as 3):
- Acute renal failure (only exclude if reason for dialysis)
- Diagnosis of HIV
- Diagnosis of sickle cell disease
- Nephritic/nephrotic syndromes
- Chronic glomerulonephritis
- Autoimmune/vasculitic nephropathy
- Congenital/familial (inc APKD) nephropathies

B. **Hypertension:**

- **Inclusion criteria** (if any present mark as 1, if none present mark as 0):
- Documented diagnosis of hypertension
- Evidence of current or past elevated SBP (>140mmHg) and/or DBP (>90mmHg)
- Evidence of current or past treatment for hypertension

C. **Type 2 Diabetes:**

- **Inclusion criteria** (if any present mark as 1, if none present mark as 0):
- Documented diagnosis of type 2 diabetes
- Evidence of current or past treatment for type 2 diabetes
- HbA1C ≥6.5% ever

6. Re-append column B to reviewers mark sheet and calculate sensitivity, specificity, PPV and NPV. Initial statistics will be based on a ‘perfect match’ only (ex: B12 = CKD: 1, HTN: 1, T2D: 0). The table below translates the algorithm status to the equivalent validation status.

| **Algorithm Status** | **CKD** | **HTN** | **T2D** |
| --- | --- | --- | --- |
| D1 | 1 | 0 | 1 |
| DH1 | 1 | 1 | 1 |
| H1 | 1 | 1 | 0 |
| C1 | 0 | 0 | 1 |
| C2 | 0 | 1 | 1 |
| C3 | 0 | 1 | 0 |
