## Supplementary Note 2 for "Deep learning on electronic medical records identifies distinct subphenotypes of diabetic kidney disease driven by genetic variations in the *Rho* pathway"

**B1. BIOBANK PATIENTS**

**N = 10852**

**LEGEND**

All counts reflect output of given box as a subset of parent box where applicable (no individual counts for given box alone)

P1 = Path 1 total

P2 = Path 2 total => B2 + B6 + B7

P3 = Path 3 total => B2 + B7

P4 = Path 4 total => B2 + B8 + B11

P5 = Path 5 total => B5 + B12 + B11

!P4 = B8 – B11

!P5 = B12 – B11

Further detail about the paths is given in Note 7 in the following pages

**Identifying Type 2 Diabetes Cases**

**B12. Treated with T2 Diabetes medication?**

**B2. Patients diagnosed with T2 Diabetes**

**Note 3**

**B7. Treated with T2 Diabetes medication**

Y

N

**EXCLUDE**

**B3. Patients diagnosed with T1 Diabetes**

**EXCLUDE**

**B8. No Diabetes medication or insulin**

**B4. Patients diagnosed with T1 or T2 Diabetes**

**Note 1**

**Note 2**

**N = 785**

**N = 3257**

**B2 υ B3**

**N = 3294**

**B10. Has T1 Diabetes diagnosis OR < 2 T2 diagnosis dates?**

**B11 (A&B). hemoglobin A1C > 6.5%**

**B13. Type 2 Diabetes Patients**

**B5. Patients NOT Diagnosed with Diabetes**

**B6. Patients treated with insulin at any time**

**B9. Never taken T2 Diabetes medication**

Y

Y

N

N

**Note 4**

**B1 MINUS B4**

**B6 MINUS B7**

**N = 7558**

**Note 5**

**Note 6**

**DISTINCT TOTAL = 2476**

**N = 1911**

**N = 305**

**P1 = 331**

**P2 = 1275**

**P3=1836**

**N = 636**

**P4 = 299**

**P5 = 10**

**N = 81**

**N = 7477**

**!P4=486**

**!P5=71**

**B2 MINUS (B6 υ B7)**

**N = 785**

**Note 1.** All T2 Diabetes ICD-9 Codes:

-250.00, 250.10, 250.20, 250.30, 250.40, 250.50, 250.60, 250.70, 250.80, 250.90, 250.02, 250.12, 250.22, 250.32, 250.42, 250.52, 250.62, 250.72, 250.82, 250.92

ICD-10 codes :

E11,E11.0,E11.00,E11.01,E11.2,E11.21,E11.22,E11.29,E11.3,E11.31,E11.311,E11.319,E11.31,E11.321,E11.329,E11.33,E11.331,E11.339,E11.34,

E11.341,E11.349,E11.35,E11.351,E11.359,E11.36,E11.39,E11.4,E11.40,E11.41,E11.42,E11.43,E11.44,E11.49,E11.5,E11.51,E11.52,E11.59,E11.6,

E11.61,E11.610,E11.618,E11.62,E11.620,E11.621,E11.622,E11.628,E11.63,E11.630,E11.638,E11.64,E11.641,E11.649,E11.65,E11.69,E11.8,E11.9

**Note 2.** All T1 Diabetes ICD-9 Codes:

-250.01, 250.11, 250.21, 250.31, 250.41, 250.51, 250.61, 250.71, 250.81, 250.91, 250.03, 250.13, 250.23, 250.33, 250.43, 250.53, 250.63, 250.73, 250.83, 250.93

ICD-10 codes :

E10,E10.1,E10.10,E10.11,E10.2,E10.21,E10.22,E10.29,E10.3,E10.31,E10.311,E10.319,E10.32,E10.321,E10.329,E10.33,E10.331,E10.339,E10.34,

E10.341,E10.349,E10.35,E10.351,E10.359,E10.36,E10.39,E10.4,E10.40,E10.41,E10.42,E10.43,E10.44,E10.49,E10.5,E10.51,E10.52,E10.59,E10.6,

E10.61,E10.610,E10.618,E10.62,E10.620,E10.621,E10.622,E10.628,E10.63,E10.630,E10.638,E10.64,E10.641,E10.649,E10.65,E10.69,E10.8,E10.9

**Note 3.** Including all drugs with active ingredient insulin, list of terms used to search for associated patients:

-“APIDRA”, “APIDRA SOLOSTAR”, “EXUBERA”, “HUMALOG”, “HUMALOG KWIKPEN”, “HUMALOG MIX 50/50”, “HUMALOG MIX 50/50 KWIKPEN”, “HUMALOG MIX 50/50 PEN”, “HUMALOG MIX 75/25”, “HUMALOG MIX 75/25 KWIKPEN”, “HUMALOG MIX 75/25 PEN”, “HUMALOG PEN”, “HUMULIN 50/50”, “HUMULIN 70/30”, “HUMULIN 70/30 PEN”, “HUMULIN BR”, “HUMULIN L”, “HUMULIN N”, “HUMULIN R”, “HUMULIN R PEN”, “HUMULIN U”, “ILETIN I”, “ILETIN II”, “INSULATARD NPH HUMAN”, “INSULIN”, “INSULIN INSULATARD NPH NORDISK”, “INSULIN NORDISK MIXTARD (PORK)”, “LANTUS”, “LENTARD”, “LENTE”, “LENTE ILETIN II”, “LENTE ILETIN II (PORK)”, “LENTE INSULIN”, “LEVEMIR”, “LEVEMIR”, “MIXTARD HUMAN 70/30”, “NOVOLIN 70/30”, “NOVOLIN L”, “NOVOLIN N”, “NOVOLIN R”, “NOVOLOG”, “NOVOLOG MIX 50/50”, “NOVOLOG MIX 70/30”, “NPH ILETIN I (BEEF-PORK)”, “NPH ILETIN II”, “NPH ILETIN II (PORK)”, “NPH INSULIN”, “NPH PURIFIED PORK ISOPHANE INSULIN”, “PROTAMINE ZINC & ILETIN I (BEEF-PORK)”, “PROTAMINE ZINC AND ILETIN II”, “PROTAMINE ZINC AND ILETIN II (PORK)”, “PROTAMINE ZINC INSULIN”, “REGULAR ILETIN II”, “REGULAR ILETIN II (PORK)”, “REGULAR INSULIN”, “REGULAR PURIFIED PORK INSULIN”, “SEMILENTE”, “SEMILENTE INSULIN”, “ULTRALENTE”, “ULTRALENTE INSULIN”, “VELOSULIN”, “VELOSULIN BR”, “VELOSULIN BR HUMAN”

**Note 4.** There is no way at present for the lab to know if tests ordered as “Glucose” were ‘fasting’ glucose or ‘random’ glucose tests. Therefore we captured all patients that had any glucose test with a result > 225 mg/dl.

"GLUCOSE (POCT)", "GLUCOSE", "WB GLUCOSE - VEN", "WB GLUCOSE-VEN (POCT)", "WB GLUCOSE-ART (POCT)", "WB GLUCOSE - ART", "GLUCOSE	FASTING"

**Note 5.** List of terms used to search for all Type 2 Diabetes medications:

"ACARBOSE", "ACTOPLUS MET", "ACTOPLUS MET XR", "ACTOS", "AMARYL", "AVANDAMET", "AVANDARYL", "AVANDIA", "DIABETA", "DUETACT", "FORTAMET", "GLIMEPIRIDE", "GLIPIZIDE", "GLIPIZIDE AND METFORMIN HYDROCHLORIDE", "GLIPIZIDE AND METFORMIN HYDROCHLORIDE", "GLUCOPHAGE", "GLUCOPHAGE XR", "GLUCOTROL", "GLUCOTROL XL", "GLUCOVANCE", "GLUMETZA", "GLYBURIDE", "GLYBURIDE (MICRONIZED)", "GLYBURIDE AND METFORMIN HYDROCHLORIDE", "GLYNASE", "JANUMET", "METAGLIP", "METFORMIN HYDROCHLORIDE", "MICRONASE", "PIOGLITAZONE", "PRANDIMET", "PRANDIN", "PRECOSE", "REPAGLINIDE", "RIOMET", "ROSIGLITAZONE", "ROSIGLITAZONE MALEATE", "ROSIGLITAZONE MALEATE; GLIMEPIRIDE", "ROSIGLITAZONE MALEATE; METFORMIN HYDROCHLORIDE"

**Note 6.** To be excluded from having ‘< 2 T2 diagnosis dates’, patients must have 2 DISTINCT dates or more, that is not 2 diagnoses on the same day and not including null date values.

**Note 7.** The gray colored counts in the flowchart that are labeled beginning with “P=…” as opposed to “N=…” are the path counts. These path counts are special as the union of these 5 sets together will give the final count of distinct type 2 diabetes patients at the end of the flowchart. Those counts that begin with “!P=…” indicate patients that were excluded in the B11 condition element before reaching the end of the flowchart. The path naming scheme is also followed in the accompanying SQL code.
